# Maternal SARS-CoV-2 impacts fetal placental macrophage programs and placenta-derived microglial models of neurodevelopment

**DOI:** 10.1101/2023.12.29.23300544

**Authors:** Lydia L. Shook, Rebecca A. Batorsky, Rose M. De Guzman, Liam T. McCrea, Sara M. Brigida, Joy E. Horng, Steven D. Sheridan, Olha Kholod, Aidan M. Cook, Jonathan Z. Li, Brittany A. Goods, Roy H. Perlis, Andrea G. Edlow

**Affiliations:** Vincent Center for Reproductive Biology, Massachusetts General Hospital, Boston, MA, USA; Department of Obstetrics, Gynecology and Reproductive Biology, Harvard Medical School, Boston, MA, USA; Data Intensive Studies Center, Tufts University, Boston, MA, USA; Center for Genomic Medicine, Massachusetts General Hospital, Boston, MA, USA; Department of Psychiatry, Massachusetts General Hospital, Harvard Medical School, Boston, MA, USA; Department of Medicine, Brigham and Women’s Hospital, Harvard Medical School, Boston, MA, USA; Thayer School of Engineering and Program, Dartmouth College, Hanover, NH, USA; Department of Molecular and Systems Biology, Geisel School of Medicine, Dartmouth College, Lebanon, NH, USA

**Author notes:** Corresponding Author: Andrea G. Edlow, MD, MSc Vincent Center for Reproductive Biology Massachusetts General Hospital 55 Fruit Street, Thier Research Building, 903B, Boston, MA 02114. Equal Contributions (Lydia L. Shook, Rebecca A. Batorsky, Brittany A. Goods, Roy H. Perlis, Andrea G. Edlow). **Disclosure Statement:** R.H.P. has received fees for service as a scientific advisor to Belle Artificial Intelligence, Burrage Capital, Circular Genomics, Genomind, Swan Artificial Intelligence Studios, and Vault Health. A.G.E. serves as a consultant for Mirvie, Inc. and receives research funding from Merck Pharmaceuticals outside of this work. The other authors have no disclosures to report.

**Keywords:** Hofbauer cells, microglia, single-cell RNA sequencing, fetal brain, placenta, neurodevelopment, neuroimmune, SARS-CoV-2, COVID-19

## Abstract

The SARS-CoV-2 virus activates maternal and placental immune responses, which in the setting of other infections occurring during pregnancy are known to impact fetal brain development. The effects of maternal immune activation on neurodevelopment are mediated at least in part by fetal brain microglia. However, microglia are inaccessible for direct analysis, and there are no validated non-invasive surrogate models to evaluate *in utero* microglial priming and function. We have previously demonstrated shared transcriptional programs between microglia and Hofbauer cells (HBCs, or fetal placental macrophages) in mouse models. Here, we assessed the impact of maternal SARS-CoV-2 on HBCs isolated from term placentas using single-cell RNA-sequencing. We demonstrated that HBC subpopulations exhibit distinct cellular programs, with specific subpopulations differentially impacted by SARS-CoV-2. Assessment of differentially expressed genes implied impaired phagocytosis, a key function of both HBCs and microglia, in some subclusters. Leveraging previously validated models of microglial synaptic pruning, we showed that HBCs isolated from placentas of SARS-CoV-2 positive pregnancies can be transdifferentiated into microglia-like cells, with altered morphology and impaired synaptic pruning behavior compared to HBC models from negative controls. These findings suggest that HBCs isolated at birth can be used to create personalized cellular models of offspring microglial programming.

## INTRODUCTION

Multiple population-based studies have suggested that maternal infection during pregnancy may have a transgenerational impact on offspring neurodevelopment. Initial work found that the incidence of schizophrenia was increased after influenza pandemics in Finland (1), Denmark (2), and the UK (3). Subsequent registry studies directly examining the association of maternal infection requiring hospitalization during pregnancy with diagnoses of autism and other neurodevelopmental disorders in offspring also found risk to be increased (4, 5). Using electronic health records, we identified an increased risk of delayed acquisition of speech and motor milestones, beyond that attributable to prematurity, in a US cohort of offspring whose mothers had SARS-CoV-2 during pregnancy. (6, 7). Similarly, authors of a prospective cohort study of 127 children in Brazil found an increased risk of neurodevelopmental delay with *in utero* SARS-CoV-2 exposure during pregnancy (8), and a recent meta-analysis of smaller studies identified additional evidence of neurodevelopmental sequelae – including reductions in fine motor and problem-solving skills – in infants with *in utero* SARS-CoV-2 exposure compared to unexposed and pre-pandemic cohorts (9). If these early signals foreshadow an increased risk of neurodevelopmental disorders in childhood and adulthood, the public health implications could be profound, given the significant number of pregnancies exposed to SARS-CoV-2 infection.

Despite the convergence of studies suggesting that maternal viral infection may increase offspring risk for neurodevelopmental disorders, the precise biological mechanisms leading to offspring neurodevelopmental vulnerability are not known. Direct placental and fetal infection with SARS-CoV-2 virus is uncommon based on current evidence (10–13), and thus vertical transmission is unlikely to be a major cause of neurodevelopmental sequelae. Animal models of maternal immune activation (MIA), in which offspring of pregnant dams treated with an immune stimulus recapitulate the behavioral hallmarks of human neurodevelopmental disorders, have been used for decades to investigate candidate *in utero* mechanisms of neurodevelopmental programming (14–17). Embryonic microglia have emerged as central mediators of offspring neuropathology in the setting of MIA (14). However, microglia from surviving offspring are inaccessible for direct analysis in humans, necessitating alternative models for evaluating the impact of SARS-CoV-2 on the fetal brain.

Prior work from our group has identified remarkable similarities in the transcriptional programs and reactivity of fetal placental macrophages, or Hofbauer cells (HBCs), and fetal brain microglia isolated from mouse embryos (18, 19). These two cell types share an embryonic origin in the fetal yolk sac (20, 21), and both carry the imprint of the in utero environment, with fetal yolk sac-derived macrophages serving as the progenitors for the lifelong pool of microglia (22, 23). Here, we investigate the impact of SARS-CoV-2 exposure on the transcriptional profiles of HBC subpopulations to gain insight into fetal resident tissue macrophage programming. Our results demonstrate that HBCs are a heterogeneous cell type, with eight subpopulations exhibiting distinct cellular programs, and that maternal SARS-CoV-2 infection is associated with varying impact on function in these subpopulations. Assessment of differentially expressed genes implies impaired phagocytosis in specific subclusters, a key function of both HBCs and microglia; we confirm these effects using a previously validated assay of microglial synaptic pruning via synaptosome phagocytosis. In aggregate, we demonstrate the application of HBC-based cellular models to gain non-invasive insight into the impact of *in utero* exposures on fetal brain development.

## RESULTS

### Hofbauer cells are a heterogeneous population with subclusters demonstrating both M1- and M2-like transcriptional signatures

Placental chorionic villous tissues were collected from N=12 birthing individuals: N=4 from individuals who had a positive SARS-CoV-2 nasopharyngeal PCR test during pregnancy, and N=8 from birthing individuals with a negative PCR at delivery and no history of a positive SARS-CoV-2 test during pregnancy. In SARS-CoV-2 positive maternal cases, infections occurred remote from delivery (median 19.5 weeks). No participants had received a COVID-19 vaccine prior to or during pregnancy and no placental samples were infected with SARS-CoV-2 at delivery (defined as having detectable SARS-CoV-2 viral load in a validated assay sensitive to 40 copies/mL) (24, 25). Thus, SARS-CoV-2 positive cases were defined by maternal infection in pregnancy, not placental or fetal infection. Additional participant characteristics are provided in Table 1.

**Table 1.**
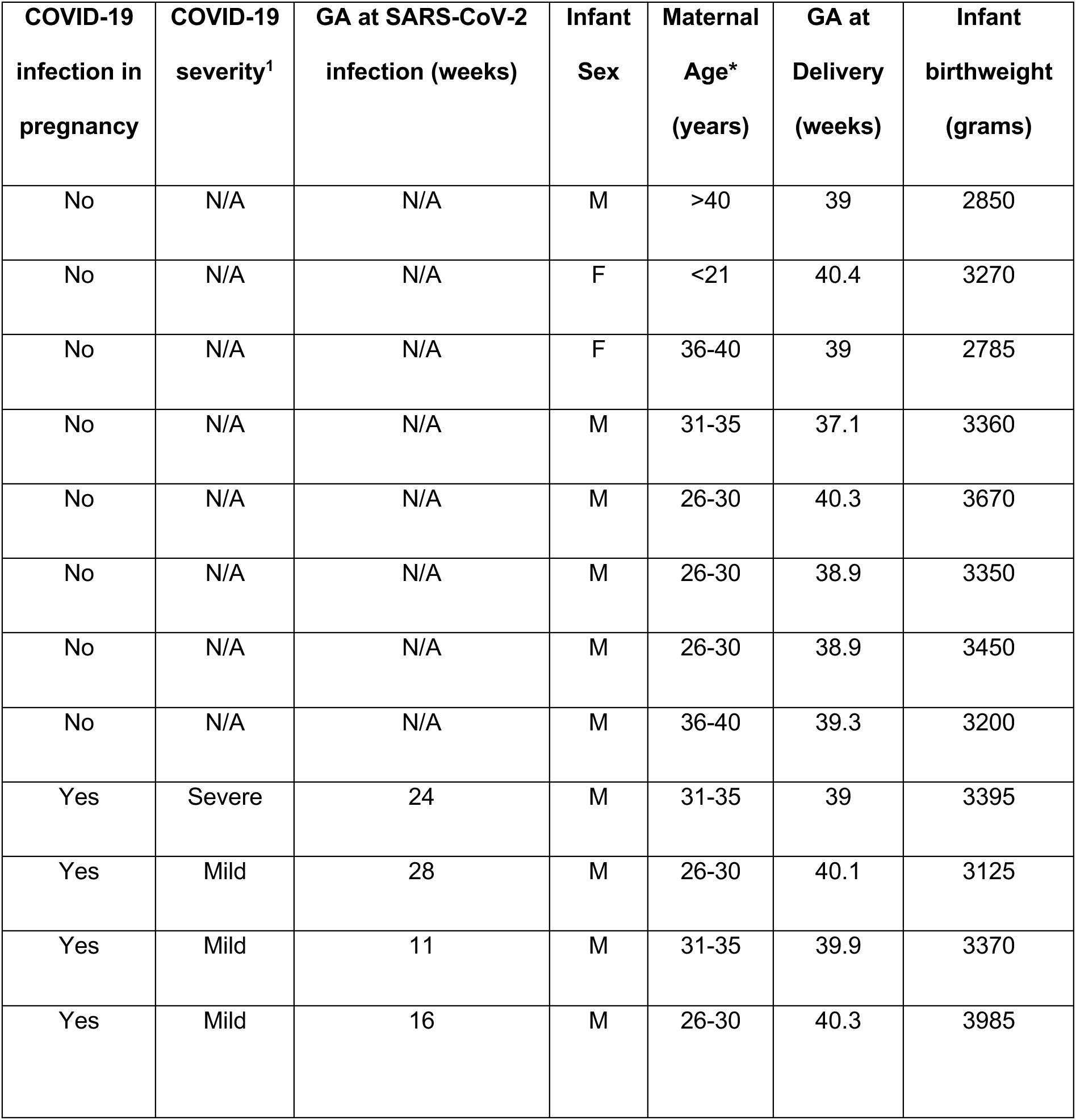
Clinical information of study participants. GA: Gestational age. M: male. F: female. N/A: not applicable. No participants had received a COVID-19 vaccine prior to delivery. ^1^COVID-19 severity was defined by NIH criteria. *Maternal age is provided as a range to preserve participant anonymity.

To assess the HBC transcriptome, we first used a previously-described protocol to obtain primarily HBCs from placental villi; in this protocol, a Percoll-based gradient and negative bead-based selection steps are used to isolate putative HBCs from other cell types present in the chorionic villi (including trophoblasts, fibroblasts) (26). Single-cell RNA sequencing was then performed on all cell suspensions (10x Genomics). After quality control filtering to remove putative doublets and cells with less than 300 identified genes, we obtained a dataset comprised of a total of 70,817 cells. We then performed sample integration and graph-based clustering to identify broad cell types (Figure S1A). Based on analyses of marker gene expression (Figure S1B), we found that the majority of cells in our dataset had marker gene expression consistent with monocytes/macrophages, and that other cell types were represented to a lesser extent, including some fibroblasts, vascular endothelial cells (VECs), extravillous trophoblasts (EVTs), leukocytes (NK cells, CD8+ T cells, B cells), neutrophils and red blood cells (Figure S1C). From this dataset, we excluded all cell types that were not identifiable as macrophages/monocytes. After additional quality control filtering for nUMIs, gene counts, and percent mitochondrial reads (see Methods), this resulted in a dataset containing 31,719 high-quality placental macrophages/monocytes. All subsequent analyses were performed with this final dataset.

After re-processing selected cells for quality control as described, we identified 10 total subclusters of macrophages/monocytes (Figure 1A), with representation of each subcluster across donors from both SARS-CoV-2 positive cases and controls (Figure S2A). To distinguish HBCs, which are placental macrophages of fetal origin, from macrophages or monocytes of maternal origin, we used cells isolated from male placentas (N=10). Male fetal origin was confirmed by high expression of *DDX3Y* and low expression of *XIST* in 8 subclusters; these were labeled HBC 0-7 (Figure 1B). The macrophage cluster with high expression of *XIST* (consistent with maternal origin) was annotated as placenta-associated maternal macrophages and monocytes (PAMMs, Figure 1B) (27). A small cluster of monocytes – identified as such by high expression of monocyte marker genes *S100A8*, *S100A9*, and *TIMP1* – demonstrated equal expression levels of both *DDX3Y* and *XIST*, suggesting that both fetal and maternal cells were present in this monocyte cluster. To further support HBC cluster annotation, we next compared the overall gene expression profiles of each cluster to a previously published single-cell dataset derived from human first-trimester placenta and decidua (28). In this analysis, all putative HBC subclusters showed highest correlation with HBC expression profiles from this dataset, whereas the monocyte and PAMM clusters had higher correlation with decidual macrophages than HBCs (Figure 1C).

**Figure 1.**
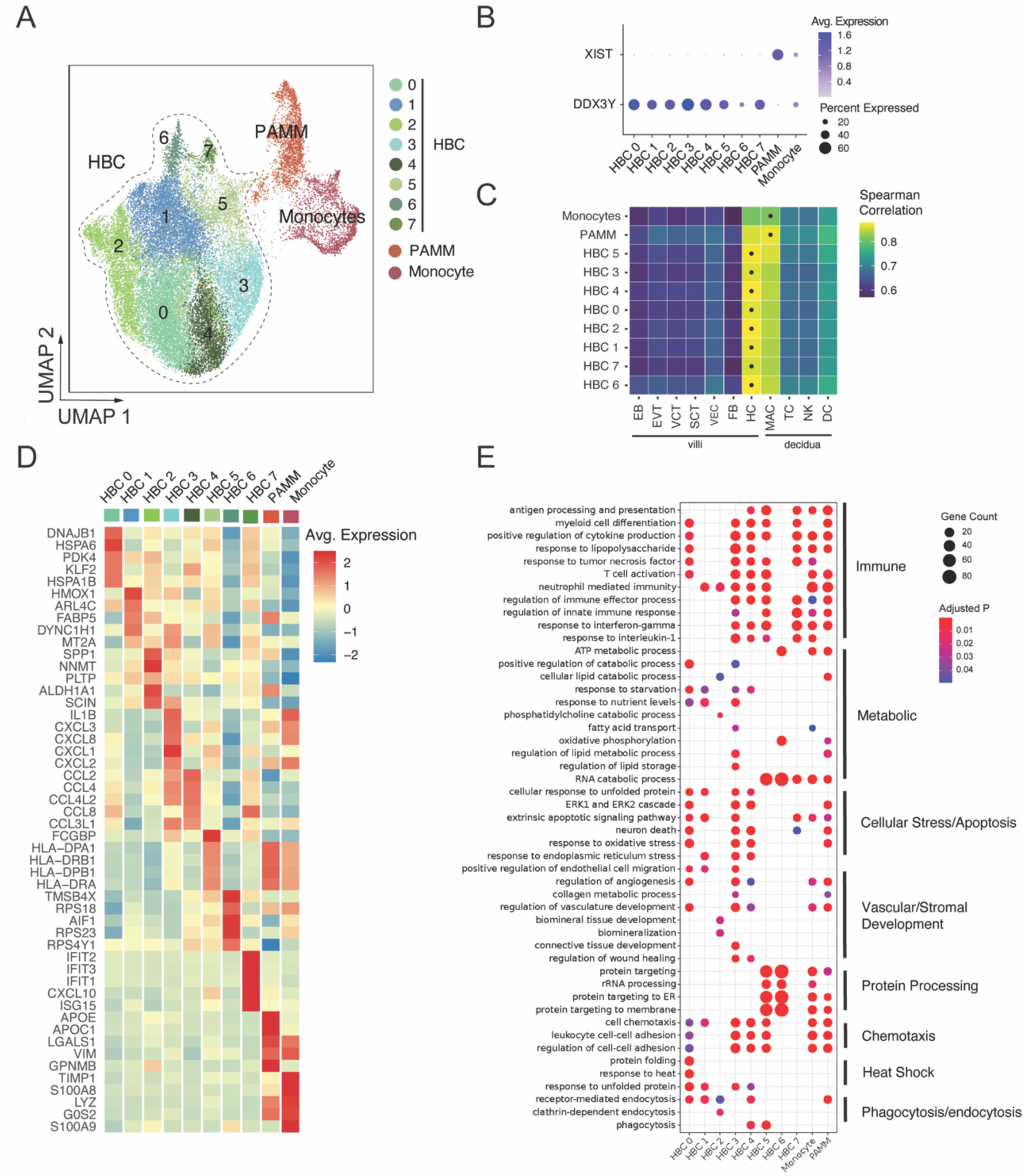
Transcriptomic profiles of fetal and maternal macrophages and monocytes isolated from term placentas with and without SARS-CoV-2 infection during pregnancy. (**A**) Uniform Manifold Approximation and Projection (UMAP) visualization of 31,719 high-quality placental macrophage and monocyte cells enriched from placentas of pregnancies with (N=4) and without (N=8) SARS-CoV-2 infection shows 10 clusters. HBC: Hofbauer cell; PAMM: placenta-associated maternal monocyte/macrophage. (**B**) Cluster-specific expression of *DDX3Y*, expressed only in fetal cells, and *XIST*, expressed only in maternal cells, in placentas from individuals carrying a male fetus (N=10). (**C**) Correlation of cluster-average gene expression with annotated cell types identified by Suryawanshi et al., *Sci Adv,* 2018. Each heatmap shows Spearman correlation coefficients. Highest correlation coefficient per cluster is indicated by black dots. HBC clusters were most highly correlated with Suryawanshi HBC clusters, PAMM cluster most correlated with decidual macrophages. (**D**) Heatmap displaying expression (log_2_ fold change) of the top 5 marker genes per cluster. (**E**) Gene Ontology (GO) Biological Process enrichment analysis for cluster marker genes. GO terms displayed were curated from the top significant GO terms in each cluster, selecting the processes most relevant to macrophage function, and reducing redundancy. Gene Count gives the number of genes in the query set that are annotated by the relevant GO category. GO terms with an adjusted p-value < 0.05 were considered significantly enriched.

To delineate differences in the identity and functions of HBC subclusters, we next assessed top marker genes defining each subcluster, shown as an average heatmap (Figure 1D). Marker genes for HBC clusters 3, 4, 5, and 7 suggested involvement in classic M1 macrophage/pro-inflammatory activities. HBC cluster 3 demonstrated high expression of chemokine (C-X-C motif) ligand genes (*CXCL*s) as well as pro-inflammatory marker genes *IL1B*, *IL1A*, *TNF*, and *NFKB1*. HBC cluster 4 demonstrated high expression of multiple CC chemokine ligand genes (CCLs) in a profile similar to that observed in HBCs responding to lipopolysaccharide stimulation *in vitro* (29). HBC cluster 5 was characterized by high expression of genes encoding major histocompatibility complex (MHC) class II molecules (human leukocyte antigen (HLA)-DRA/B1 and -DP) and Fc-gamma binding protein (FCGBP), suggesting a role in antigen presentation to CD4+ T cells. MHC class II molecules are critical to antigen-specific responses, and upregulation of HLA complexes and antigen presentation pathways has been observed in proteomic analyses of HBCs stimulated with the viral dsRNA mimic poly(I:C) (29). HBC cluster 7 demonstrated marker genes from the interferon-induced protein with tetratricopeptide repeats (*IFIT*) family (*IFIT2*, *IFIT3*), *CXCL10*, *ISG15*, and *MX1*, associated with a pro-inflammatory type 1 interferon antiviral response. To gain further insight into the biological processes reflected in each HBC subcluster, we performed Gene Ontology (GO) enrichment analyses of cluster marker genes (Figure 1E). As would be expected given marker gene expression noted previously, pathways involved in M1-like immune/inflammatory responses were enriched in HBC 3, 4, 5, and 7, including “response to interleukin-1,” “response to interferon-gamma,” “response to tumor necrosis factor,” and “positive regulation of cytokine production.”

Gene signatures of HBC clusters 0, 1, and 2 reflected engagement in specific stress response processes, particularly response to inflammation and/or tissue damage, to ultimately support placental function. HBC 0 and HBC 1 were characterized by genes encoding heat shock proteins and other proteins involved in endoplasmic reticulum stress and the unfolded protein response, such as *HSPA6, HSPA1B, DNAJB1, HSP90B1*, *HSPA5* and *BAG3*. The unfolded protein response represents a homeostatic response to restore balance when endoplasmic reticulum stress is sensed and to modulate and/or resolve inflammation (30). Additionally, HBC 0’s high expression of *PDK4* and *KLF2* may suggest involvement in attenuating oxidative stress responses and reducing pro-inflammatory cytokine production (31, 32), and HBC 1’s high expression of *FABP5* and *HMOX1* suggests engagement in anti-inflammatory responses against heme-induced toxicity and induction towards M2 polarization (33, 34). GO enrichment analysis of these clusters similarly demonstrated enrichment of pathways such as “response to unfolded protein”, “response to heat”, “response to endoplasmic reticulum stress”, and pathways related to cellular stress response and apoptosis (e.g. “extrinsic apoptotic signaling pathway”, “response to oxidative stress” and “ERK1 and ERK2 cascade”).

GO enrichment analysis also suggested both HBC 0 and HBC 1 were engaged in homeostatic functions including “receptor-mediated endocytosis”, “regulation of angiogenesis” and “vascular development”, and nutrient-sensing functions such as “response to nutrient levels” and “response to starvation.” HBC 2 was characterized by high expression of the genes encoding secreted phosphoprotein 1 (*SPP1*) and Nicotinamide N -methyltransferase (*NNMT*), both associated with M2 (anti-inflammatory) macrophage polarization in the context of tumor-associated macrophages (35, 36); SPP1, also known as osteopontin, is secreted by HBCs and plays an important role in endothelial biology and angiogenesis (37). GO analysis of HBC cluster 2 also demonstrated enrichment in “receptor-mediated endocytosis” (involved in intracellular transport of macromolecules), as well as “cellular lipid catabolic process,” and processes associated with stromal tissue development.

HBC 6 was characterized by expression of genes involved in regulation of actin polymerization and cytoskeleton organization (*TMSB4X* and *AIF1*, which encodes the canonical microglial marker Iba1 (38)) and several ribosomal proteins including *RPS18, RPS23,* and *RPS4Y1*. GO enrichment analysis of this cluster demonstrates highly specific enrichment of protein processing pathways (e.g. “protein targeting” pathways, “cytoplasmic translation,” “translational initiation”) and pathways related to RNA catabolism, oxidative phosphorylation, and ATP metabolism. Marker genes of the maternal PAMM subcluster included *APOE*, *APOC1*, *VIM, LGALS1*, and *GPNMB* among others, an expression pattern consistent with previously reported maternal placental macrophage transcriptional profiles (39, 40). High expression of *LGALS1/3* and *GPNMB* by the PAMM cluster suggests a role in inflammation regulation (41–43), which was echoed by GO analyses identifying enrichment in immune response and immunomodulatory pathways (e.g. “antigen processing and presentation, “positive regulation of cytokine production”, and “regulation of innate immune response”) . In addition, GO enrichment analysis demonstrated PAMM were engaged in lipid metabolic processes and receptor-mediated endocytosis, consistent with their known involvement in lipid engulfment, and tissue repair/scar formation (37, 39).

### Maternal SARS-CoV-2 infection drives cluster-specific differences in immune signaling and metabolic pathways

Once the baseline functions of HBC and PAMM subclusters had been established, we then sought to characterize the impact of maternal SARS-CoV-2 infection on the transcriptomic profile of HBC subclusters. To do so, we identified differentially expressed genes (DEG) by maternal SARS-CoV-2 status within each cluster. DEG were defined using log fold-change threshold of 0.2 and adjusted p-value of 0.05 (see Methods for full details). We first verified that each subcluster included representation from both SARS-CoV-2+ cases and controls (Figure 2A, top panel). The proportion of cells from cases versus controls was consistent across all subclusters, except for HBC 0, which demonstrated a relatively high contribution of control donor cells (Figure S2A). Of the 8 HBC clusters, a majority (5) were significantly impacted by maternal SARS-CoV-2 infection: HBC 0, 1, 2, 3, and 5 (Figure 2A, bottom panel). In contrast, HBC clusters 4, 6, and 7 had very few DEG in the setting of maternal SARS-CoV-2 exposure, with three, zero, and two DEG respectively. Of the 5 highly impacted clusters, HBC 1 and HBC 5 had the highest number of DEG by maternal SARS-CoV-2, with 723 and 566 DEG, respectively. PAMMs were impacted by SARS-CoV-2 to a lesser extent, with 67 DEG identified. Both up- and down-regulated DEG were identified across all impacted clusters.

**Figure 2.**
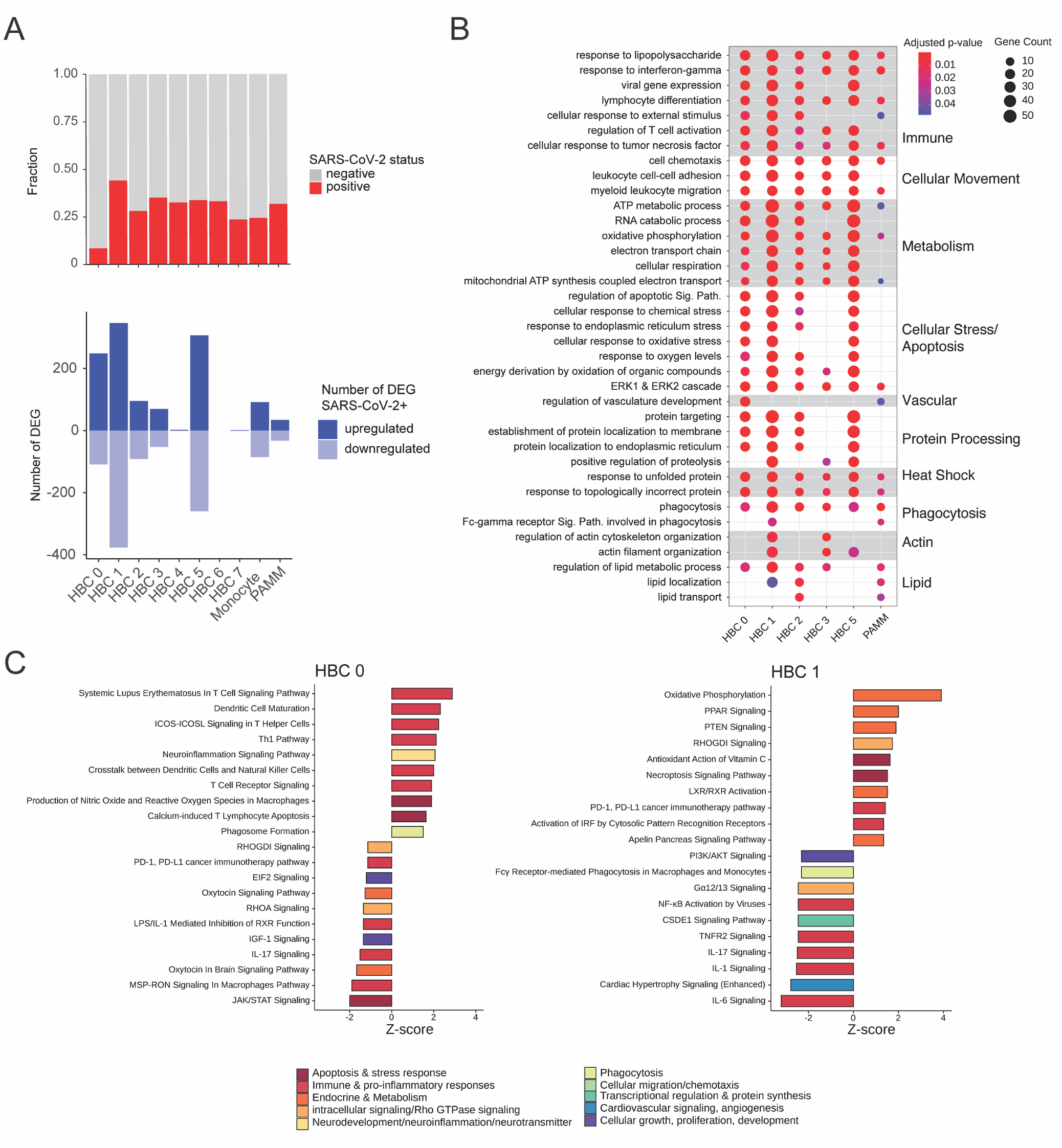
Impact of maternal SARS-CoV-2 infection on Hofbauer cell subclusters. DEG: differentially expressed genes. (**A**) Barplot demonstrating proportion of cells per cluster from SARS-CoV-2 positive cases (red) and negative controls (gray), top panel. Number of DEG upregulated (dark blue) and downregulated (light blue) by SARS-CoV-2 exposure per cluster, bottom panel. (**B**) Gene Ontology (GO) Biological Process enrichment analysis for DEG. Gene Count gives the number of genes in the query set that are annotated by the relevant GO category. GO terms with an adjusted p-value < 0.05 were considered significantly enriched. (**C**) Ingenuity Pathway Analysis (IPA) of DEG for HBC clusters 0 (left) and 1 (right). Canonical pathways with absolute Z-score ≥1 and adjusted p-value < 0.05 are shown. IPA analysis for remaining HBC clusters depicted in Supplement.

GO pathway enrichment analysis of DEG indicated that in the context of maternal SARS-CoV-2 infection, specific pathways involved in immune responses were enriched in all impacted HBC clusters (HBC 0, 1, 2, 3 and 5), including response to lipopolysaccharide, response to interferon-gamma, cellular response to tumor necrosis factor, and regulation of T cell activation (Figure 2B). Additionally, all subclusters were enriched for pathways related to cellular movement, such as cell chemotaxis, leukocyte cell-cell adhesion, and myeloid leukocyte migration; heat shock-related pathways (unfolded protein response); and phagocytosis pathways.

GO enrichment analysis also indicated some biological processes that were only dysregulated in specific clusters in the setting of maternal SARS-CoV-2 infection (Figure 2B). For example, regulation of vascular development was only dysregulated in HBC 0 and the PAMM cluster, regulation of lipid metabolism and transport were dysregulated in all clusters except HBC 5, more cellular energy utilization pathways (e.g. ATP metabolism, electron transport chain/oxidative phosphorylation and cellular respiration) and cellular stress/apoptosis pathways were impacted in HBC clusters relative to the PAMM cluster, and protein processing and actin cytoskeleton organization pathways were only dysregulated in HBC clusters but not PAMMs. Taken together, these functional analyses suggest that in the context of maternal SARS-CoV-2 infection, HBC subclusters and PAMMs are differentially impacted, with key dysregulated biological processes including innate immune and pro-inflammatory signaling, cell chemotaxis and migration, cellular ATP and lipid metabolism, cellular phagocytosis, and the unfolded protein response.

To better understand the impact of SARS-CoV-2 on HBC functions, we next used Ingenuity Pathway Analysis (IPA), which predicts strength and directionality (i.e. activation or suppression) of enriched canonical pathways by subcluster. In this analysis, pathways with absolute Z-score value greater than 1 (consistent with IPA being able to “call” a direction of dysregulation of the pathway) and Benjamini Hochburg-adjusted p<0.05 were included and displayed by subcluster (Figure 2C and Figure S2B-F). Z-scores ≥ 1 indicate upregulated signaling in the pathway and ≤ 1 indicate downregulated pathway signaling (44). Both HBC 1 and HBC 2 subclusters exhibited a primarily anti-inflammatory response to SARS-CoV-2, with activation of PPAR signaling and oxidative phosphorylation, a metabolic profile associated with an anti-inflammatory/pro-resolution phase macrophage signature (45, 46). In HBC 1 and 2, suppression of IL-6, IL-1, and IL-17 pathways, and activation of LXR/RXR signaling pathways in SARS-CoV-2+ cases suggests involvement in resolution of inflammation, as LXR/RXR pathway activation in macrophages is associated with inhibition of inflammatory gene expression and promotion of lipid metabolism (47). Also consistent with an anti-inflammatory role, HBC 2 showed strong suppression of the Coronavirus Pathogenesis Pathway and activation of Oxytocin Signaling Pathway, the latter of which is involved in attenuating oxidative and cellular inflammatory responses in macrophages (48).

Conversely, HBC 0 and 3 demonstrated primarily activated pro-inflammatory immune responses in SARS-CoV-2+ cases, with increases in LPS/IL-1 mediated inhibition of RXR (HBC 3), Interferon induction (HBC 3), Neuroinflammation signaling (HBC 0 and 3), T-cell signaling (HBC 0 and 3), and Production of nitric oxide and reactive oxygen species (HBC 0). Metabolic processes were suppressed in both clusters, including Oxytocin signaling pathway (HBC 0), Sirtuin signaling (HBC 3), and MSP-RON signaling (HBC 0 and 3) (48–50). In the context of maternal SARS-CoV-2 infection, subcluster HBC 5 presented a mixed picture of both pro- and anti-inflammatory signaling, with upregulation of interferon, EIF 2, neuroinflammation and T cell related signaling pathways, balanced by upregulation of anti-inflammatory pathways such as PPAR signaling and downregulation of pro-inflammatory signaling pathways such as Coronavirus Pathogenesis pathway, FAK and TNF-mediated signaling pathways.

Compared to HBC subclusters, PAMMs were less impacted overall by maternal SARS-CoV-2 at a transcriptomic level, with 67 DEG identified. In the setting of maternal SARS-CoV-2 infection, PAMMs showed activation of pathways involved in immune responses including Production of Nitric Oxide and Reactive Oxygen Species, B-cell signaling pathways, Interferon induction, and activation of the pattern recognition receptor TREM-1 signaling. Similar pro-inflammatory patterns were observed for monocytes, including activation of antiviral response pathways and Th1 signaling pathways, and suppression of MSP-RON signaling (Figure S2C). Taken together, these analyses point to transcriptional shifts in some but not all subclusters in response to SARS-CoV-2, with a greater response by HBCs compared to PAMMs, driven by a combination of immune activation/pro-inflammatory signature in subclusters HBC 0 and HBC 3 and an anti-inflammatory tissue repair signature in clusters HBC 1 and HBC 2.

### Maternal SARS-CoV-2 infection impacts HBC transcriptional programs associated with phagocytosis, neuroinflammation, and neurological disorders

Tissue-resident macrophages promote resolution of inflammation through phagocytosis of apoptotic cells, invading pathogens, or cellular debris (51, 52). Phagocytosis is also a key function of microglia in early brain development (53–55). IPA functional analysis of SARS-CoV-2-specific HBC signatures demonstrated that macrophage phagocytosis (Figure 3A) and neurological disease-related pathways (Figure 3B) were key functions and pathways implicated by the cluster-specific gene expression signatures. Figure 3 summarizes the impact of maternal SARS-CoV-2 infection on placental macrophage phagocytosis (Figure 3A, 3C), illustrating the potential for altered HBC gene programs to provide insight into both fetal brain microglial function and the impact of maternal SARS-CoV-2 infection on neurodevelopment (Figure 3B, 3D). These analyses predicted SARS-CoV-2-associated suppression of phagocyte chemotaxis and cell movement pathways (e.g. reduced “activation of phagocytes”, “recruitment of phagocytes”, “cell movement of phagocytes”, “adhesion of phagocytes”) in HBC 1, 2 and 5, consistent with the suppression of synaptosome phagocytosis (a proxy for synaptic pruning) observed in subsequent experiments using in vitro Hofbauer cell induced microglial assays, detailed below. In contrast to the consistent suppression of phagocytosis in HBC clusters 1, 2 and 5, HBC clusters 0 and 3 and the PAMM cluster demonstrated activation of phagocytosis-related pathways including “Phagocytosis” (HBC 0), “Immune response of phagocytes” (HBC 0), “Phagocytosis by macrophages” (HBC 3) and “Cellular infiltration by phagocytes” (HBC 3). A representative phagocytosis pathway from IPA and expression of its constituent genes by cluster is depicted as a heatmap in Figure 3C. Cluster-specific alterations in phagocyte movement in the setting of maternal SARS-CoV-2 infection were primarily driven by expression differences in *CXCL2*, *NFKB1A*, *NFKB1Z*, *IL1B*, *CXCL8*, *CD36*, and *ICAM1* by cluster (Figure 3C). Concordant with patterns observed in canonical pathways analyses, HBC 1 and 2 (and to a lesser extent HBC 5), which showed primarily immunomodulatory signatures, also show suppressed phagocytosis and phagocytic movement pathways, versus proinflammatory clusters HBC 0 and 3, which demonstrate activation of phagocytosis.

**Figure 3.**
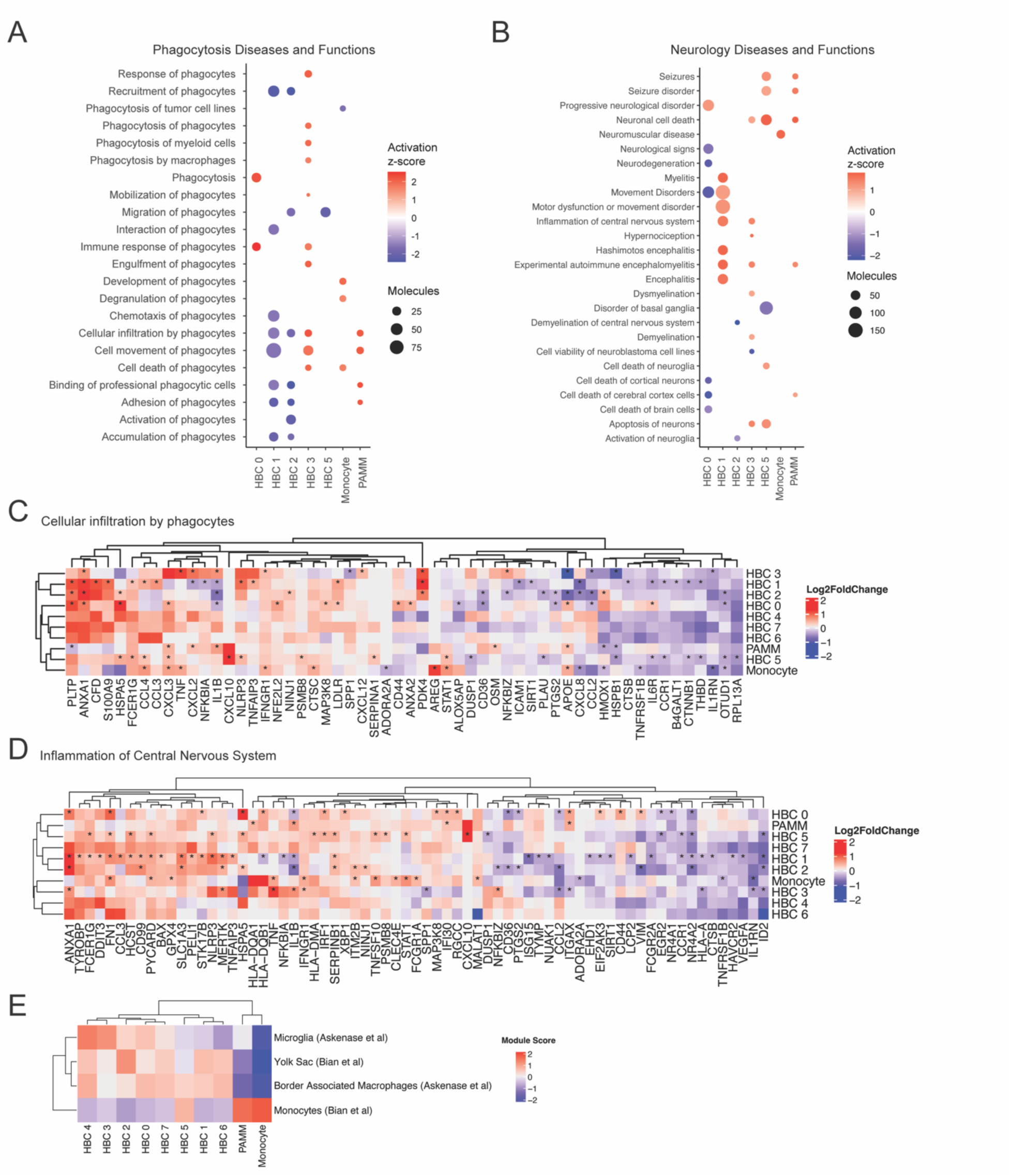
Impact of maternal SARS-CoV-2 on HBC gene programs associated with phagocytosis and neurologic disease. (**A-B**) Ingenuity Pathway Analysis (IPA) phagocytosis diseases and functions pathways (**A**), and neurologic diseases or functions (**B**), enriched for ≥ 3 DEGs, with absolute Z-score ≥1 and adjusted P-value < 0.05. Activation Z-score represented by color and number of DEGs by circle size, with red color indicating pathway activation and blue color indicating suppression. (**C-D**). Heatmap of gene expression in “Cellular Infiltration by Phagocytes” IPA Pathway (**C**) and “Inflammation of Central Nervous System” (**D**) by cluster. Color represents gene expression level (log_2_ fold change), *adjusted P-value < 0.05. (**E**). Module score by subcluster in comparison to cluster-specific gene expression of single-cell datasets from human brain: Microglia and Border Associated Macrophages (Askenase et al., *Sci Immunol*, 2021) and Yolk Sac Macrophages and Monocytes (Bian et al., *Nature*, 2020). Color indicates module score.

In addition to phagocytosis, pathways relevant to neurologic disease and microglial functions emerged as key dysregulated pathways in the setting of maternal SARS-CoV-2 infection. We therefore assessed whether DEG of HBC subclusters map to neuroinflammatory/neurodevelopmental pathways and functions in IPA, and plotted pathway activation Z-scores by subcluster (Figure 3B). The transcriptional signature of HBC 1 – in which Fc-gamma receptor-mediated phagocytosis (Figure 2C) and other previously-described phagocytosis pathways (Figure 3A) are suppressed in SARS-CoV-2+ cases – is also associated with increased neuroinflammation, including positive activation Z-scores for “Inflammation of central nervous system”, “Myelitis”, and “Encephalitis.” Cluster-specific expression of the genes in the “Inflammation of central nervous system pathway” is depicted in Figure 3D, with upregulation of signaling in this pathway driven by increased expression of *ANXA1*, *FN1*, *CCL3*, *SLC1A3, NLRP3,* and

*MERTK*, among others. Interestingly, HBC 3 – in which “Inflammation of Central Nervous System” is also predicted to be activated after maternal SARS-CoV-2 infection and whose transcriptional signature is consistent with activation in phagocytosis pathways, also implicates increased “Apoptosis of neurons” and “Neuronal cell death” (Figure 3B). In a developmental context, this pattern may represent a functional rather than pathologic gene signature in response to SARS-CoV-2, as microglia (resident brain macrophages) play a key role in neuronal cell turnover, regulation of neural progenitors, and synaptic rewiring in early neurodevelopment, all via phagocytosis (56). Thus, increased phagocytosis by tissue-resident macrophages might be an adaptive response to SARS-CoV-2-associated inflammation, while reduced phagocytosis could be a pathologic or maladaptive response to maternal immune activation (e.g., reduced microglial phagocytosis and reduced synaptic pruning associated with maternal immune activation is thought to be a key aspect of the pathogenesis of autism (57–59)). Taken together, these data support the notion that HBC transcriptional signatures provide insight into protective versus pathologic microglial programming in the setting of an immune challenge such as SARS-CoV-2.

Prior work from our group in a mouse model has shown that HBCs and fetal brain microglia share transcriptional profiles and responses to maternal obesity, an immune-activating exposure (19). To further probe the potential connection between transcriptional signatures of HBC subclusters and brain microglia in humans, we next compared marker genes from HBC subclusters with gene modules from published human single cell atlases of macrophages derived from adult and embryonic brain (Figure 3E) (60, 61). Nearly all HBC subclusters scored highly for gene signatures found in microglia, yolk sac macrophages, or CNS border associated macrophages, compared with monocytes and PAMMs. HBC 2 and 1 exhibited greatest similarity to yolk sac macrophages whereas HBC 3 and 4 were most like microglia isolated from adult brain samples. In contrast, monocytes and PAMMs were most similar to circulating monocytes, which is concordant with their shared myeloid lineage (37). This analysis supports the concept that HBCs isolated from full term human placenta share transcriptional signatures with yolk sac macrophages and fetal brain microglia, and thus may offer insights into global reprogramming of fetal macrophage populations, including those of the fetal brain, in the setting of maternal immune-activating exposures.

### HBCs isolated from placentas of SARS-CoV-2 positive pregnancies can be transdifferentiated to microglia-like cells (HBC-iMGs)

To gain insight into the functional consequences of SARS-CoV-2 exposure on HBC populations, we used a previously-validated model in which HBCs isolated from SARS-CoV-2 positive cases (N=4) and a subset of SARS-CoV-2 negative controls (N=4) were cultured in media containing IL-34 and GM-CSF to obtain transdifferentiated microglia-like cells (HBC-iMGs), as we have previously described (see Methods) (62–64). Following culture, we assessed the expression of multiple markers associated with microglial identity, including IBA1, TMEM119, PU.1, and P2RY12 (65, 66), and identified expression of all markers in the majority of HBC-iMGs from both SARS-CoV-2 positive cases and negative controls (Figure 4A, Figure S3A-B).

**Figure 4.**
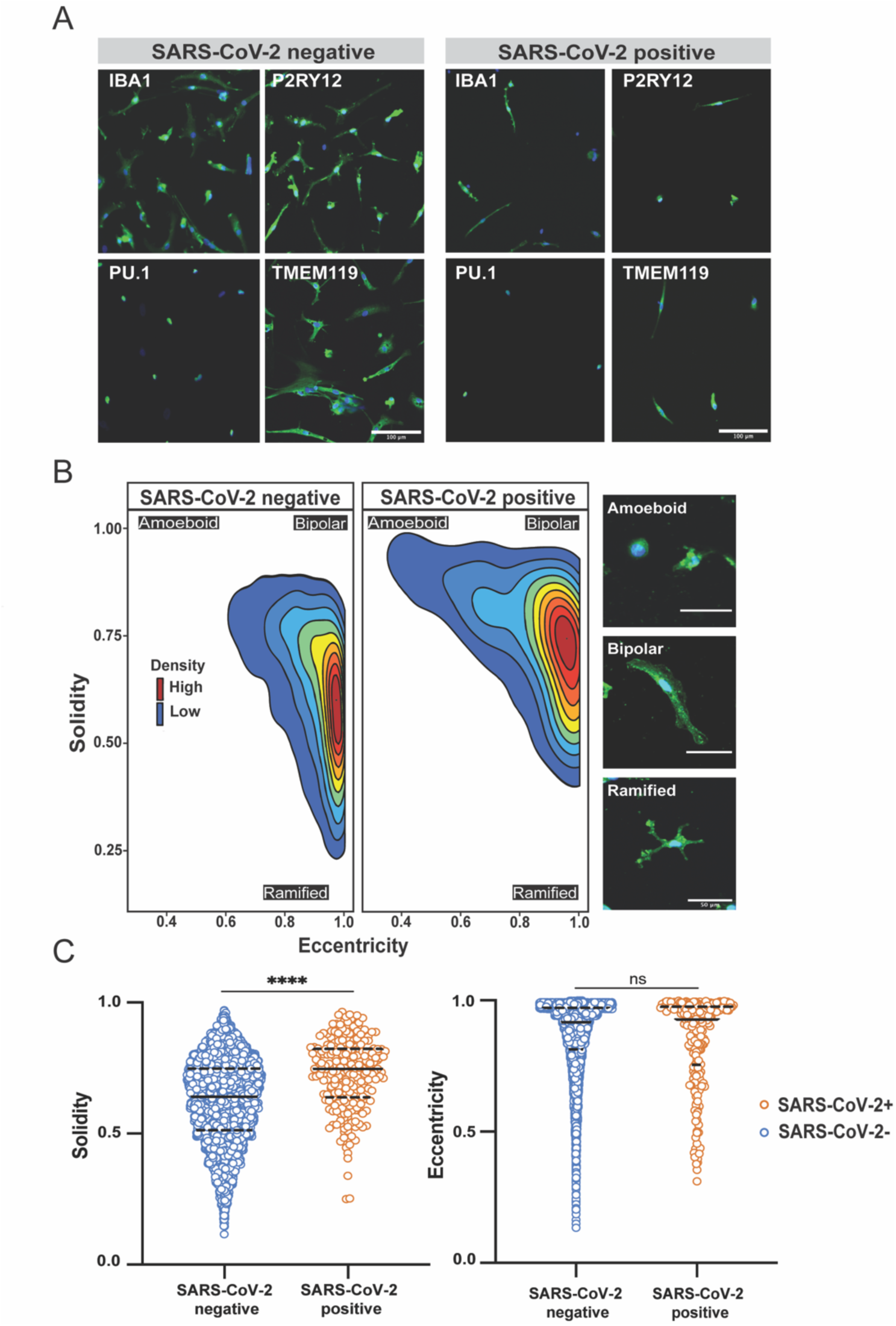
Phenotypic characterization of HBC-iMGs by direct cytokine reprogramming. HBC-iMGs: Hofbauer cells transdifferentiated toward microglia-like cells. (**A**) Images of HBC-iMGs from SARS-CoV-2 positive cases (n=4) and negative controls (n=4), immunostained for microglial markers: IBA1, PU.1, P2RY12, TMEM119. Scale bar = 100 µm. (**B**) Morphology-smoothed density plots (solidity vs. eccentricity) for SARS-CoV-2 negative and positive samples as indicated. Cells from negative controls exhibit a more ramified morphology than those from positive cases, suggestive of a less activated phenotype. Red color shows high density and blue is low density. Representative confocal microscopy images of amoeboid, bipolar, and ramified HBC-iMGs. Scale bar = 50 µm. (**C**) Violin plots represent distribution of cell solidity (left) and eccentricity (right) measurements from SARS-CoV-2 negative controls (blue, n=5223 cells) and positive cases (orange, n=237 cells). Solid lines represent median values and dashed lines interquartile range. Group differences assessed by Mann-Whitney U Test. ****P<0.0001. ns = not significant.

#### SARS-CoV-2-exposed HBC-iMGs demonstrate increased amoeboid morphology and impaired phagocytic behavior compared to HBC-iMGs from uninfected control placentas

To assess differences in cell phenotype by SARS-CoV-2 exposure, we first evaluated cellular morphology of IBA1-positive HBC-iMGs by quantitative assessment of two morphological characteristics: eccentricity (amoeboid vs bipolar shape) and solidity (amoeboid/bipolar vs ramified shape) (Figure 4B-C). In this analysis, HBC-iMGs from SARS-CoV-2 negative controls demonstrated a more ramified morphology than those from positive cases, indicated by their lower solidity and high eccentricity (Figure 4B-C). More ramified microglial morphology is generally typical of resting-state, tissue-surveilling microglia in vivo (67, 68). In contrast, a greater proportion of HBC-iMGs generated from SARS-CoV-2 positive cases demonstrated higher solidity and smaller cell size (Figure 4B-C, Figure S3C), consistent with a more amoeboid appearance. While this morphology is classically attributed to an immune-activated state (69), it is also typical of microglial patterns observed in fetal states (67, 70).

Transcriptional analyses of HBC clusters pointed to a cluster-specific impact of maternal SARS-CoV-2 on phagocytosis pathways. HBC clusters with the greatest similarity to embryonic/yolk sac microglia (e.g. HBC 1, 2) also exhibited cellular programs suggestive of impaired phagocytosis. Using a previously-validated model of synaptic pruning (62–64), a key physiologic function of microglia in early brain development, we next tested the functional capability of HBC-iMGs to engage in phagocytosis. In this assay, HBC-iMGs were co-cultured for 3 hours with pHrodo Red-labeled neuronal synaptosomes derived from human induced pluripotent stem cells prior to fixation and imaging. This pH-sensitive label fluoresces following intracellular engulfment (see Methods). Synaptosome engulfment by IBA1-positive cells was then measured by quantifying fluorescence using confocal microscopy images with CellProfiler software applied for segmentation and thresholding (Figure 5A). Compared to SARS-CoV-2 negative cases, HBC-iMGs from positive cases demonstrated significant impairment in synaptosome phagocytosis, reflected by a reduced phagocytic index (Figure 5B). Phagocytic index was reduced across all SARS-CoV-2 positive samples, and was driven by reduced phagocytic uptake per cell, regardless of the proportion of cells engaged in phagocytosis in any given sample. (Figure S3D-E). In conjunction with the morphologic changes, and consistent with the transcriptomic signatures observed in a subset of HBC, these functional phenotypes support a dysregulated activation state following SARS-CoV-2 infection.

**Figure 5.**
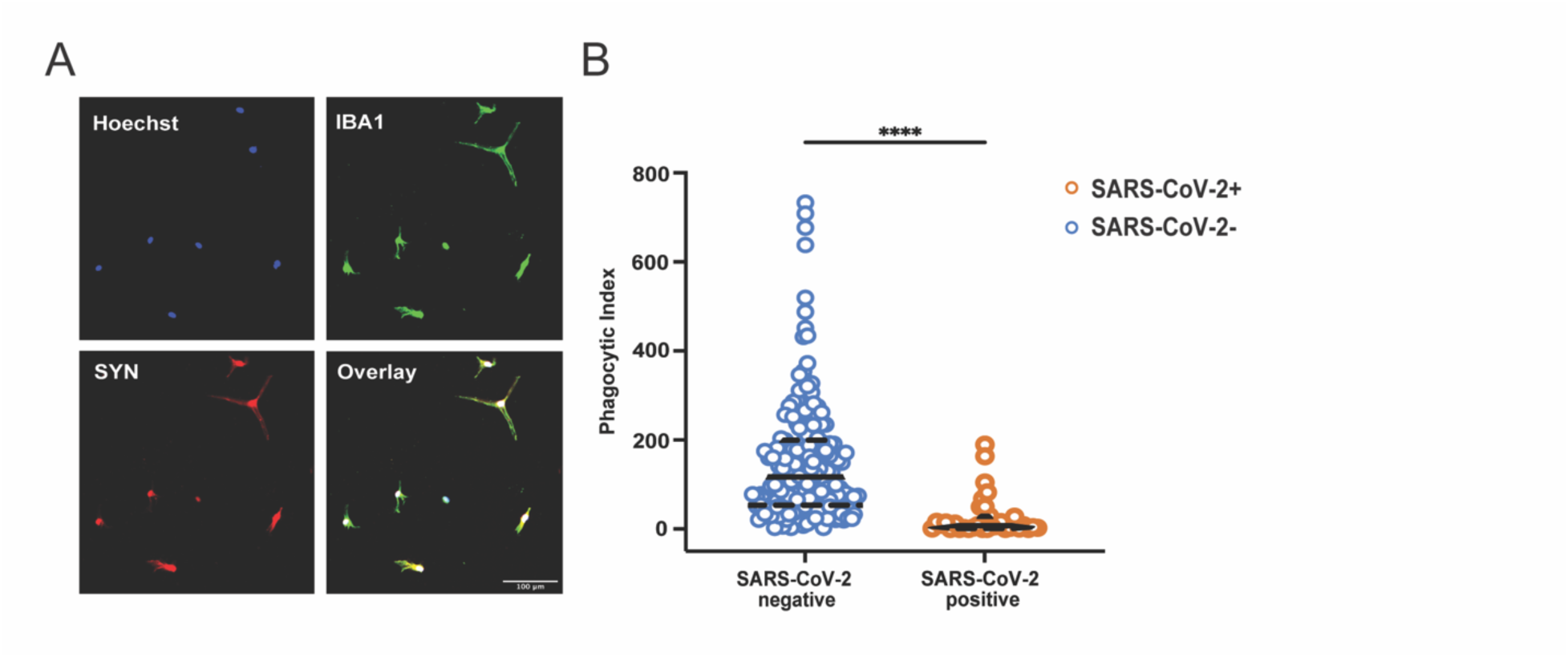
Impact of maternal SARS-CoV-2 on HBC-iMG synaptosome engulfment. HBC-iMGs: Hofbauer cells transdifferentiated toward microglia-like cells. (**A**) Representative image showing colocalization of pHrodo-red labeled synaptosomes (SYN) and IBA1 positive HBC-iMGs. Hoechst = nuclear stain. Scale bar = 100 µm. (**B**) Violin plots of phagocytic index of image fields from SARS-CoV-2 negative controls (blue, n=187 fields) and positive cases (orange, n=32 fields). Phagocytic index is calculated as synaptosome area in pixels divided by cell count per image field. Solid lines represent median values and dashed lines interquartile range. Group differences assessed by Mann-Whitney U Test. ****P<0.0001.

## DISCUSSION

Data from observational cohorts suggests an increased neurodevelopmental risk of offspring exposed *in utero* to maternal SARS-CoV-2 infection (6–8) but the underlying mechanism for offspring brain vulnerability remains unknown. Studies have consistently demonstrated that maternal SARS-CoV-2 infection drives alterations in immune cell populations and pro-inflammatory responses at the maternal-fetal interface (71–78) that have the capacity to impact the fetal brain (79). Even in the absence of direct viral transmission to the fetus, profiling of umbilical cord blood immune cell populations and the serum proteome demonstrates that maternal SARS-CoV-2 infection can shape fetal and neonatal immunity (40, 80–82). Prior bulk and single-cell transcriptomic analyses have also revealed significant reprogramming at the maternal-fetal interface following SARS-CoV-2 infection during pregnancy (40, 71, 72, 77), yet granular information on fetal placental cell populations has been relatively limited by their lower representation in these studies.

Here we report single-cell RNA-seq data that provide new insights into the heterogeneous functions that fetal placental macrophages, or Hofbauer cells, and maternal resident placental macrophages and monocytes or PAMMs, perform at baseline, and how these programs are altered in the setting of maternal SARS-CoV-2 infection. We found that maternal SARS-CoV-2 infection in pregnancy, even distant from delivery and in the absence of placental infection, was associated with significant alterations in the transcriptional programs of Hofbauer cells. These programs were more significantly impacted than those of maternal placental macrophages, as indicated by number of DEG. Effects of maternal SARS-CoV-2 infection were subcluster-specific, with phagocytosis being a key dysregulated function, and affected Hofbauer cell clusters exhibited signatures consistent with neuroinflammation and neurologic disease. We directly tested this predicted dysregulation using validated in vitro models of HBC-based induced microglia (HBC-iMGs) (62–64), confirming that SARS-CoV-2 infection altered HBC-iMG morphology and function. SARS-CoV-2 exposed HBC-iMGs were more ameboid in shape and exhibited reduced synaptosome phagocytosis, an assay that serves as a proxy for synaptic pruning. Notably, reduced synaptic pruning by microglia has been identified as a key mechanism in the pathogenesis of autism (57, 83), a neurodevelopmental disease associated with maternal immune activation and viral infection in pregnancy (59, 84, 85). Considering the shared fetal origin between Hofbauer cells and brain microglia (20, 21), this work indicates Hofbauer cells’ potential to serve as a more accessible cell type at birth that could provide information about fetal brain immune programming, which in turn could alter neurodevelopmental trajectories after in utero exposure to maternal SARS-CoV-2 infection.

Our study is unique in its in-depth, focused interrogation of fetal immune cell populations of the placenta in the context of a remote maternal viral infection. Through sex-chromosome-specific gene expression mapping, we were able to reliably assign fetal cell identity to 8 subtypes of HBCs, engaged in a myriad of functions at baseline. Similar to the work of Thomas et al. in first trimester placenta, we identified subclusters with transcriptional programs associated with angiogenesis and tissue remodeling, as well as clusters enriched for immune defense functions (39). Concordant with prior single-cell RNA sequencing studies of the placenta in the context of SARS-CoV-2 infection (40, 86), we identified that even in the absence of direct placental infection or active COVID-19 disease at the time of delivery, maternal exposure to SARS-CoV-2 remote from delivery had a profound impact on the transcriptional programs of the fetal macrophage population, and to a lesser extent maternal PAMMs.

We defined a broad range of responses to SARS-CoV-2 across HBC subclusters, including some clusters with relatively few DEG and others with significant transcriptional shifts. Impacted HBC subclusters demonstrated transcriptional programs evoking the changes observed in neuroinflammation, and the same subclusters exhibited alterations in phagocytosis and in chemotaxis and cellular movement. To investigate these results further we created induced microglial cellular models from the same samples (HBC-iMG). Phenotypic and functional analyses of HBC-iMGs from SARS-CoV-2 positive samples demonstrated a shift toward more amoeboid morphology and significant impairments in synaptosome phagocytosis. Reduced phagocytic efficiency appeared to result from reduced capacity for synaptosome uptake within the cell, rather differences in the proportion of cells engaged in phagocytosis.

We demonstrated for the first time that HBCs can be used to create microglia-like cell models, applying this approach to gain insight into fetal brain immune programming in the context of maternal SARS-CoV-2 infection. As yolk-sac derived macrophages that colonize the fetal brain early in development (20), microglia play a fundamental role in neurogenesis by promoting neural precursor cell proliferation, axonal outgrowth, and synaptic wiring throughout development (53–55). A key function of microglia in normal neurodevelopment also includes selective phagocytosis of excess neuronal precursors and synapses to edit and refine the architecture of neuronal communication (55, 87). Evidence from animal models of maternal immune activation (MIA) suggests that microglia are keenly responsive to maternal innate immune signaling, and MIA-induced disruption of normal microglial function can recapitulate social deficits and other behaviors correlative of those observed in neurodevelopmental disorders such as autism spectrum disorder and schizophrenia (57, 58, 88, 89).

A primary strength of our study is inclusion of rigorously phenotyped individuals without a history of prior SARS-CoV-2 infection or vaccination and of contemporaneously enrolled control subjects. We thus were able to examine the impact of maternal SARS-CoV-2 on an immunologically naïve cohort in the absence of prior immunity to SARS-CoV-2, with a consequence being that we could not assess the impact of prior vaccination. Neither the impact of COVID-19 severity nor fetal sex could be assessed in this study due to the study design (primarily focused on symptomatic infection and male samples as a proof of principle study) and small sample size. Sex differences will be particularly important to assess in future work, given the importance of fetal sex on offspring neurodevelopmental vulnerability and fetoplacental programming (90, 91). Taken together, our results suggest the ability of HBC-iMGs to serve as personalized cellular models of microglial programming in the setting of maternal exposures, including SARS-CoV-2 and potentially other environmental exposures that might impact neurodevelopment. They demonstrate potential mechanisms by which these exposures may contribute to adverse neurodevelopmental outcomes.

## METHODS

### Study design and participant enrollment

In this study, 12 pregnant individuals with full-term, singleton pregnancies delivering at Massachusetts General Hospital (March 2021 - August 2021) were included. Participants were classified as SARS-CoV-2 positive (N=4) if they had symptomatic COVID-19 infection during pregnancy, confirmed by positive SARS-CoV-2 nasopharyngeal PCR test. Participants were classified as SARS-CoV-2 negative (n=8) if they did not have a positive SARS-CoV-2 nasopharyngeal PCR or COVID-19 symptoms during pregnancy and had a negative SARS-CoV-2 nasopharyngeal PCR at delivery upon universal COVID-19 screening on Labor and Delivery. Pregnant individuals were eligible for inclusion if they were 18 years or older and were delivering during the COVID-19 pandemic. For this study, individuals with prior COVID-19 vaccination were excluded. A study questionnaire and review of the electronic health record was used to determine key demographic variables such as maternal age, gestational age at delivery, gestational age at positive COVID-19 test, COVID-19 disease severity at diagnosis, any prior diagnoses of COVID-19 or history of COVID-19 vaccination, and infant sex and birthweight.

### Placenta collection and processing

Placentas were obtained within 20 minutes after delivery and submerged in Cytowash media (Dulbecco’s Modified Eagle Medium (DMEM) containing 2.5% FBS, 1% Penicillin-Streptomycin, 0.1% Gentamicin) and stored at 4°C until cell isolation. Isolation of Hofbauer cells was performed using previously described protocols (26); reagents are listed in Supplemental Table S1 and isolation workflow and study procedures are depicted in Supplemental Figure S4. Briefly, placental chorionic villi were separated from fetal membranes and decidua, washed in DPBS wash, and mechanically homogenized. Placental tissue was then serially digested in Collagenase Digestion Buffer, Trypsin Digestion Buffer, and Collagenase Digestion Buffer 2. Undigested tissue was removed by passage through sterile gauze and 100uM filter. The cell suspension was centrifuged at 257g for 8 min at 4°C, washed, spun again, and resuspended in media. The cells were then suspended in 4mL of 20% Percoll and 5mL of 35% Percoll was underlayed through a #1 glass Pasteur pipette (92). After centrifugation for 30 minutes at 4°C without brake at 1000g, cells were isolated from the interphase layer, washed in media, and spun at 257g for 8 minutes at 4°C. Cell pellets were immunopurified by negative selection by incubation with anti-EGFR (to remove syncytiotrophoblasts) and anti-CD10 (to remove fibroblasts) conjugated to magnetic Dynabeads, prepared as previously described (76), for 20 minutes at 4°C. Tubes were placed on a DynaMagTM magnet for 5 minutes to magnetically bind cytotrophoblasts (anti-EGFR) and fibroblasts (anti-CD10) – allowing media containing unbound placental macrophages to pass through into collection tubes. Cells were cryopreserved in 90% FBS and 10% dimethyl sulfoxide (DMSO) at 1-10 million cells/vial and stored at -80°C for downstream analyses. SARS-CoV-2 viral loads were assessed in all placental tissues using qPCR as previously described, with 40 copies/mL as limit of detection (24, 25).

### Single-cell RNA-sequencing (scRNA-Seq) data analysis

#### Sequencing

Cryopreserved HBCs were thawed at 37°C and diluted with RPMI 1640 including 10% FBS and 1% Pen/Strep. The cell suspension was centrifuged at 300g for 5 min at room temperature, with the brake off. The supernatant was aspirated and the cell pellet was resuspended in media. Dead cells were removed using OptiPrepTM Density Gradient Medium (Sigma) and cell count and viability of cells were calculated using LunaFX7 automated counter. Cells were immediately loaded onto 10x Genomics platform with a loading target of approximately 10,000 viable cells/sample. Libraries were sequenced on an Illumina NextSeq 2000 P3 flowcell machine with a sequencing target of 25,000 reads per cell.

#### Initial cluster identification

Raw reads were aligned to reference genome GRCh38 and quantified using Cell Ranger (version 6.0.1, 10x Genomics) and after initial cellranger filtering an average of 6,295 cell/sample and 20,459 reads/cell were present. Putative doublet cells were removed using predictions generated from DoubletFinder (v2.0.3) as were cells containing less than 300 identified genes, which resulted in an object containing 70,817 cells. All samples were integrated to remove batch effects from individuals using the Seurat Single Cell Transform workflow (Seurat version 4.3.0) with the top 2,000 variable features. Cells were clustered using the Louvain algorithm on the shared nearest neighbor graph and visualized by UMAP using the first 30 principal components. Several clustering resolutions were used to scan through the data to identify a resolution (0.3) that allowed us to identify top-level cell types based on marker genes. Marker genes were identified using the Wilcoxon rank sum test with the following parameters: only.pos = TRUE, min.pct = 0.2, logfc.threshold = 0.5. Additionally, expression of well-known cell-type markers was assessed to refine top-level identities: hofbauer, fibroblasts, NK cells/CD8 T cells, VECs (vascular endothelial cells), EVTs (extravillous trophoblasts), RBCs (red blood cells), B cells, and neutrophils (Supplemental Figure S1). For subsequent analyses, we created a subset of the data including only cells identified as “Hofbauer,” which included macrophage and monocyte populations.

#### Subcluster analysis

The data were re-integrated and processed similarly as described above to identify macrophage/monocyte subclusters. Only high-quality cells were retained (mitoRatio < 0.25 and nUMI > 1000 and < 9681, or 3 standard deviations above the mean UMI count). The number of subclusters was optimized by iteratively clustering across several cluster resolutions, and identifying the resolution that provided non-redundant clusters (resolution = 0.3) as determined by marker gene identification with Seurat’s Wilconcon rank-sum test (only.pos = TRUE, min.pct = 0.3, and logfc.threshold = 0.5). Subclusters were then assigned as HBCs (0-7), PAMMs or Monocytes based on marker genes. To delineate fetal from maternal origin of subclusters, we evaluated sex-specific markers using only cells from placentas with a male fetus (N=10). This allowed for maternal vs fetal cell differentiation, as fetal cells would be expected to have increased expression of the male-specific Y-linked gene *DEAD-Box Helicase 3 Y-Linked* (*DDX3Y*) and maternal cells would exhibit high expression of the female-specific gene *X-inactive specific transcript* (*XIST*). The macrophage cluster with high expression of *XIST* consistent with maternal origin was annotated as maternal placenta-associated macrophages and monocytes (PAMMs). To further support cell cluster annotation, we also compared the overall gene expression profiles of each cluster to a previously published single-cell dataset derived from human first-trimester placenta and decidua (28).

#### Differential gene expression by SARS-CoV-2 status

For differential gene expression analysis between cells from SARS-CoV-2 positive cases and negative controls, we used the Seurat FindMarkers() within each cluster with the following parameters: test.use = “MAST”, min.pct =0.3, logfc.threshold = 0.2, latent.vars = “donor”. Genes with Benjamini-Hochberg adjusted p-value < 0.05 were considered significant.

#### Functional enrichment analyses

Gene Ontology (GO) Biological Process enrichment analysis was performed on both cluster marker genes and SARS-CoV-2 differentially expressed genes in each cluster using R package clusterProfler (v. 3.18.1) (93) and underlying database AnnotationDb org.Hs.eg.db (v3.12.0). GO terms were considered significant with adjusted p-value < 0.05. IPA Canonical Pathway and Diseases and Functions analysis were performed with IPA (Content Version: 90348151) with pathways considered significant with adjusted p-value <0.05.

### Derivation of Hofbauer cells transdifferentiated toward microglia-like cells (HBC-iMGs) by direct cytokine reprogramming

HBC-iMGs were derived from HBCs using previously described methods (62–64), with modifications as noted. Briefly, thawed HBCs were plated on Geltrex-coated 24-well plates (1 × 10^6^ cells in 0.5 mL per well) or 96-well plates (2 x 10^5^ cells in 0.1mL per well) depending on cell availability. After cells were incubated at 37°C for 24 h, the media was completely replaced with RPMI 1640 including GlutaMAX, 1% penicillin–streptomycin, 100 ng/mL of human recombinant IL-34 (Peprotech), and 10 ng/mL of GM-CSF (Peprotech). At day 6 of transdifferentiation, the cultures were assayed and subsequently fixed with 4% PFA to perform endpoint analysis using immunocytochemistry.

### HBC-iMG Immunocytochemistry

HBC-iMGs were washed twice with PBS and blocked for 1 h with 5% FBS and 0.3% Triton-X (Sigma Aldrich) in PBS at room temperature. Next, they were washed three times with 1% FBS in PBS and incubated with primary antibodies in 5% FBS and 0.1% Triton-X overnight at 4°C (Anti-IBA1, 1:500; Abcam #ab5076; Anti-TMEM119, 1:500; Abcam #ab18537; Anti-CX3CR1, 1:100, Abcam #ab8021; Anti-PU.1, 1:1000, Abcam #ab183327, and Anti-P2RY12, 1:100, Alomone Labs). Cells were then washed three times with 1% FBS in PBS and incubated in secondary antibodies (Invitrogen Alexa Fluor, 1:500) and Hoechst 33342 (1:5000) in 5% FBS and 0.1% Triton-X in PBS for 45 min at 4°C. Cells were washed two final times and imaged using the IN Cell Analyzer 6000 (Cytiva). Marker characterization was analyzed using CellProfiler (94). Cells were segmented using one of the four microglia markers used and percent marker positive cells calculated by dividing the number of marker positive cells by the number of identified nuclei, per image. A total of 12 20x images per sample were analyzed.

### Synaptosome derivation and phagocytosis assay

#### Synaptosome generation from neural progenitor cell cultures

Induced pluripotent stem cells were reprogrammed from fibroblasts and used to derive expandable neural progenitor cells, and large-scale differentiated neural cultures, as previously described (62–64). After media removal, neural cultures were collected by scraping in 10ml per T1000 flask 1× gradient buffer (ice-cold 0.32 M sucrose, 600 mg/L Tris, 1 mM NaH3CO3, 1 mM EDTA, pH 7.4 with added HALT protease inhibitor—Thermo-Fisher Scientific # 78442) and homogenized using a Dounce Tissue Grinder (Wheaton #357544 15ml) with the ‘tight’ plunger. Homogenate was collected and centrifuged at 700g g for 10 min at 4°C to remove large debris. The gradient buffer was removed by aspiration and saved on ice then the pellet was resuspended in 10 mL of 1× gradient buffer and homogenization was repeated as above. The final homogenates were combined and centrifuged at 15,000g for 15 min at 4°C. This second pellet was resuspended in 12ml 1× gradient buffer and slowly added on top of a pre-formed sucrose gradient in Ultracentrifuge Tubes (Beckman Coulter Ultra-Clear #344058) containing 12ml each 1.2 M (bottom) and 0.85 M (middle) sucrose layers. The gradients were centrifuged using Ultracentrifuge swinging bucket rotor #SW32TI at 26,500 RPM (∼80,000g) for 2 h at 4°C with the brake off. The synaptosome band (in between 0.85 and 1.2 M sucrose layers) was removed using a 5-mL syringe and 19Gx1 ½″ needle, diluted with 5-fold 1x gradient buffer then centrifuged at 20,000g for 20 min at 4°C. The final pellet was resuspended in an appropriate volume of 1× gradient buffer containing 1 mg/mL bovine serum albumin (BSA) with HALT protease and phosphatase inhibitors added, aliquoted and slowly frozen at −80°C. Protein concentration was measured by BCA and enrichment of pre-synaptic (synapsin, SNAP-25) and post-synaptic (PSD-95) markers was determined by western blot analysis.

#### Phagocytosis Assay

Synaptosomes were thawed and labeled with pHrodo Red SE (Thermo-Fisher Scientific #P36600) at 1:2 (mg dye: mg synaptosome) and incubated at room temperature for 1 hour. Labeled synaptosomes were sonicated for 1 hour before adding to HBC-iMGs at 15mg total protein per well in 24-well plates, or 3mg per well in 96-well format. HBC-iMGs with synaptosomes were incubated at 37°C for three hours and then fixed with 4% PFA for 15 minutes at room temperature. Immunocytochemistry was performed to quantify phagocytosis, with images analyzed in CellProfiler (v4.2.4). HBC-iMGs were segmented as described below using IBA1 staining and phagocytic index was calculated by dividing the signal area of pHrodo Red by the number of segmented cells, per image.

### Image analysis

CellProfiler (v4.2.4) was used to identify cellular and subcellular structures in the confocal images (94). The module CorrectIlluminationCalculate and CorrectIlluminationApply were used in all channels to correct uneven illumination and uneven background. Nuclei and cell bodies were each identified using IdentifyPrimaryObjects. Specifically, pixel diameter ranges and the automatic thresholding method Otsu were applied. The module RelateObjects was used to drop structures incorrectly identified as nuclei by ensuring they were only accepted when they have a surrounding microglia-like cell. IdentifySecondaryObjects was used to more accurately outline cells around these nuclei and avoid debris. The module IdentifyTertiaryObjects identified cytoplasm by subtracting the area of the nucleus from the cell. IdentifySecondaryObjects was used with Otsu thresholding to identify and measure synaptosomes. Background red signal was eliminated by increasing the lower bounds on the automatic threshold, using reference images from Cytochalasin treatment as a positive control of diminished phagocytosis. MaskObjects was used with the cell and synaptosome objects to omit red signal from outside the cell. Overlays of the outlines of all generated image structures were created for quality check purposes using the OverlayOutlines module. Cell area, count, and signal intensity were created with MeasureObjectSizeShape, MeasureObjectIntensity, ExportToSpreadSheet, and ExportToDatabase. RStudio 2 (1.4) was used to organize the metadata exported from CellProfiler. Phagocytic index was calculated as area of Synaptosomes divided by cell count per image. As a confirmation, the integrated intensity of Synaptosome signal divided by cell area was also checked to make sure both measures corresponded. Images containing >80 cells were omitted due to procedure inaccuracy with dense fields. Outliers were excluded by calculating a phagocytic index threshold of 3 SD above the mean. Morphology data was produced using cell level metadata from CellProfiler followed by a cleaning process matching the field level dataset cleaning. Cells with an area or synaptosome area of greater than the mean plus 3 SD were omitted. In total, 12 20x images per sample were analyzed.

### Statistics

Group differences were assessed using Mann-Whitney U tests. P values less than 0.05 were considered statistically significant. Dark lines represent median and dotted lines interquartile range, unless otherwise specified. Statistical analyses were performed in GraphPad Prism (version 9.3).

## Study approval

This study was approved by the Mass General Brigham Institutional Review Board (Protocol #2020P003538). All participants provided written informed consent prior to participation.

## Data availability

Sequencing data will be made available for download on GEO upon acceptance. R code supporting the conclusions of this manuscript is made available here: https://github.com/rbatorsky/covid-placenta-edlow.

## Author Contributions

L.L.S. and R.A.B. contributed equally, and as co-first authors. A.G.E. conceived the study and, together with R.H.P., designed the experiments. Acquisition of data: L.L.S., R.A.B., R.M.D., L.T.M., S.M.B., S.D.S., J.Z.L., S.B., J.E.H., B.A.G., R.H.P., A.G.E. Analysis and interpretation of data: L.L.S., R.A.B., R.M.D., L.T.M., O.K., S.D.S., A.M.C., B.A.G., R.H.P., A.G.E. Drafting of the manuscript: L.L.S., R.A.B., R.M.D., L.T.M., A.G.E. Revising the manuscript critically for important intellectual content: L.L.S., R.A.B., R.M.D., L.T.M., S.M.B., O.K., J.E.H., S.D.S., A.M.C., J.Z.L., B.A.G., R.H.P., A.G.E. All authors have given final approval for submission.

## Acknowledgements

NIH/NICHD: 1R01HD100022-01, 3R01HD100022-02S2, and 1U19AI167899-01 to A.G.E; 1K12HD103096 to L.L.S.; NIH/NIMH: 1RF1MH132336-01 to A.G.E. and R.H.P.; NIH: 5T32HG010464 to R.M.D.; B.A.G. was supported in part by the Geisel School of Medicine at Dartmouth’s Center for Quantitative Biology by NIH/NIGMS: P20GM130454. J.Z.L. was supported by a grant from the Massachusetts Consortium for Pathogen Readiness (MassCPR).

## References

1. Mednick SA, et al. Adult schizophrenia following prenatal exposure to an influenza epidemic. Arch Gen Psychiatry. 1988;45(2):189–192.

2. Barr CE, Mednick SA, Munk-Jorgensen P. Exposure to influenza epidemics during gestation and adult schizophrenia. A 40-year study. Arch Gen Psychiatry. 1990;47(9):869–874.

3. Cooper SJ. Schizophrenia after prenatal exposure to 1957 A2 influenza epidemic. Br J Psychiatry. 1992;161:394–396.

4. Al-Haddad BJS, et al. Long-term Risk of Neuropsychiatric Disease After Exposure to Infection In Utero. JAMA Psychiatry. 2019;76(6):594–602.

5. Atladóttir HO, et al. Maternal infection requiring hospitalization during pregnancy and autism spectrum disorders. J Autism Dev Disord. 2010;40(12):1423–1430.

6. Edlow AG, et al. Sex-Specific Neurodevelopmental Outcomes Among Offspring of Mothers With SARS-CoV-2 Infection During Pregnancy. JAMA Netw Open. 2023;6(3):e234415.

7. Edlow AG, et al. Neurodevelopmental Outcomes at 1 Year in Infants of Mothers Who Tested Positive for SARS-CoV-2 During Pregnancy. JAMA Netw Open. 2022;5(6):e2215787.

8. Santos CAD, et al. Developmental impairment in children exposed during pregnancy to maternal SARS-COV2: A Brazilian cohort study. Int J Infect Dis. [published online ahead of print: December 5, 2023]; 10.1016/j.ijid.2023.12.001

9. Pinheiro GSMA, et al. Effects of intrauterine exposure to SARS-CoV-2 on infants’ development: a rapid review and meta-analysis. Eur J Pediatr. 2023;182(5):2041–2055.

10. Kotlyar AM, et al. Vertical transmission of coronavirus disease 2019: a systematic review and meta-analysis. Am J Obstet Gynecol. 2020;0(0). 10.1016/j.ajog.2020.07.049

11. Woodworth KR, et al. Birth and Infant Outcomes Following Laboratory-Confirmed SARS-CoV-2 Infection in Pregnancy - SET-NET, 16 Jurisdictions, March 29-October 14, 2020. MMWR Morb Mortal Wkly Rep. 2020;69(44):1635–1640.

12. Flaherman VJ, et al. Infant Outcomes Following Maternal Infection with SARS-CoV-2: First Report from the PRIORITY Study. Clin Infect Dis. [published online ahead of print: 2020]; 10.1093/cid/ciaa1411

13. Dumitriu D, et al. Outcomes of Neonates Born to Mothers With Severe Acute Respiratory Syndrome Coronavirus 2 Infection at a Large Medical Center in New York City. JAMA Pediatr. [published online ahead of print: October 12, 2020]; 10.1001/jamapediatrics.2020.4298

14. Otero AM, Antonson AM. At the crux of maternal immune activation: Viruses, microglia, microbes, and IL-17A. Immunol Rev. 2022;311(1):205–223.

15. Simões LR, et al. Maternal immune activation induced by lipopolysaccharide triggers immune response in pregnant mother and fetus, and induces behavioral impairment in adult rats. J Psychiatr Res. 2018;100:71–83.

16. Baines KJ, et al. Maternal Immune Activation Alters Fetal Brain Development and Enhances Proliferation of Neural Precursor Cells in Rats. Front Immunol. 2020;11:1145.

17. Careaga M, Murai T, Bauman MD. Maternal Immune Activation and Autism Spectrum Disorder: From Rodents to Nonhuman and Human Primates. Biol Psychiatry. 2017;81(5):391–401.

18. Edlow AG, et al. Placental Macrophages: A Window Into Fetal Microglial Function in Maternal Obesity. Int J Dev Neurosci. [published online ahead of print: November 20, 2018]; 10.1016/j.ijdevneu.2018.11.004

19. Batorsky R, et al. Hofbauer cells and fetal brain microglia share transcriptional profiles and responses to maternal diet-induced obesity. bioRxiv. 2023;2023.12.16.571680.

20. Ginhoux F, et al. Fate Mapping Analysis Reveals That Adult Microglia Derive from Primitive Macrophages. Science. 2010;330(6005):841–845.

21. Gomez Perdiguero E, et al. Tissue-resident macrophages originate from yolk-sac-derived erythro-myeloid progenitors. Nature. 2015;518(7540):547–551.

22. Ginhoux F, Prinz M. Origin of microglia: current concepts and past controversies. Cold Spring Harb Perspect Biol. 2015;7(8):a020537.

23. Ginhoux F, et al. Origin and differentiation of microglia. Front Cell Neurosci. 2013;7:45.

24. Fajnzylber J, et al. SARS-CoV-2 viral load is associated with increased disease severity and mortality. Nat Commun. 2020;11(1):5493.

25. Edlow AG, et al. Assessment of Maternal and Neonatal SARS-CoV-2 Viral Load, Transplacental Antibody Transfer, and Placental Pathology in Pregnancies During the COVID-19 Pandemic. JAMA Netw Open. 2020;3(12):e2030455.

26. Tang Z, et al. Isolation of Hofbauer Cells from Human Term Placentas with High Yield and Purity: ISOLATION OF PLACENTAL HOFBAUER CELLS. Am J Reprod Immunol. 10/2011;66(4):336–348.

27. Megli C, Coyne CB. Gatekeepers of the fetus: Characterization of placental macrophages. J Exp Med. 2021;218(1). 10.1084/jem.20202071

28. Suryawanshi H, et al. A single-cell survey of the human first-trimester placenta and decidua. Sci Adv. 2018;4(10):eaau4788.

29. Pantazi P, et al. Placental macrophage responses to viral and bacterial ligands and the influence of fetal sex. iScience. 2022;25(12):105653.

30. Heck TG, et al. Suppressed anti-inflammatory heat shock response in high-risk COVID-19 patients: lessons from basic research (inclusive bats), light on conceivable therapies. Clin Sci . 2020;134(15):1991–2017.

31. Gao X, et al. PDK4 Decrease Neuronal Apoptosis via Inhibiting ROS-ASK1/P38 Pathway in Early Brain Injury After Subarachnoid Hemorrhage. Antioxid Redox Signal. 2022;36(7–9):505–524.

32. Das H, et al. Kruppel-like factor 2 (KLF2) regulates proinflammatory activation of monocytes. Proc Natl Acad Sci U S A. 2006;103(17):6653–6658.

33. Vijayan V, Wagener FADTG, Immenschuh S. The macrophage heme-heme oxygenase-1 system and its role in inflammation. Biochem Pharmacol. 2018;153:159–167.

34. Hou Y, et al. FABP5 controls macrophage alternative activation and allergic asthma by selectively programming long-chain unsaturated fatty acid metabolism. Cell Rep. 2022;41(7):111668.

35. Dong B, et al. Macrophage-Related SPP1 as a Potential Biomarker for Early Lymph Node Metastasis in Lung Adenocarcinoma. Front Cell Dev Biol. 2021;9:739358.

36. Li Y, et al. Nicotinamide N -methyltransferase promotes M2 macrophage polarization by IL6 and MDSC conversion by GM-CSF in gallbladder carcinoma. Hepatology. 2023;78(5):1352–1367.

37. Thomas JR, et al. The Ontogeny and Function of Placental Macrophages. Front Immunol. 2021;12:771054.

38. Sasaki Y, et al. Iba1 is an actin-cross-linking protein in macrophages/microglia. Biochem Biophys Res Commun. 2001;286(2):292–297.

39. Thomas JR, et al. Phenotypic and functional characterization of first-trimester human placental macrophages, Hofbauer cells. J Exp Med. 2021;218(1). 10.1084/jem.20200891

40. Doratt BM, et al. Mild/asymptomatic COVID-19 in unvaccinated pregnant mothers impairs neonatal immune responses. JCI Insight. 2023;8(19). 10.1172/jci.insight.172658

41. Ripoll VM, et al. Gpnmb is induced in macrophages by IFN-gamma and lipopolysaccharide and acts as a feedback regulator of proinflammatory responses. J Immunol. 2007;178(10):6557–6566.

42. Zhang H, et al. GPNMB plays an active role in the M1/M2 balance. Tissue Cell. 2022;74:101683.

43. Yaseen H, et al. Galectin-1 Facilitates Macrophage Reprogramming and Resolution of Inflammation Through IFN-β. Front Pharmacol. 2020;11:901.

44. Tugendreich SM. Understanding biological mechanisms in transcriptomics or proteomics datasets with Ingenuity Pathway Analysis (IPA) and Analysis Match https://www.dkfz.de/genomics-proteomics/fileadmin/Ingenuity/IPA_AnalysisMatch_White_paper.pdf.

45. Abdalla HB, et al. Activation of PPAR-γ induces macrophage polarization and reduces neutrophil migration mediated by heme oxygenase 1. Int Immunopharmacol. 2020;84:106565.

46. Viola A, et al. The Metabolic Signature of Macrophage Responses. Front Immunol. 2019;10:1462.

47. Hong C, et al. Constitutive activation of LXR in macrophages regulates metabolic and inflammatory gene expression: identification of ARL7 as a direct target. J Lipid Res. 2011;52(3):531–539.

48. Szeto A, et al. Regulation of the macrophage oxytocin receptor in response to inflammation. Am J Physiol Endocrinol Metab. 2017;312(3):E183–E189.

49. Chen YQ, Fisher JH, Wang MH. Activation of the RON receptor tyrosine kinase inhibits inducible nitric oxide synthase (iNOS) expression by murine peritoneal exudate macrophages: phosphatidylinositol-3 kinase is required for RON-mediated inhibition of iNOS expression. J Immunol. 1998;161(9):4950–4959.

50. Yoshizaki T, et al. SIRT1 inhibits inflammatory pathways in macrophages and modulates insulin sensitivity. Am J Physiol Endocrinol Metab. 2010;298(3):E419–28.

51. A-Gonzalez N, et al. Phagocytosis imprints heterogeneity in tissue-resident macrophages. J Exp Med. 2017;214(5):1281–1296.

52. Mosser C-A, et al. Microglia in CNS development: Shaping the brain for the future. Prog Neurobiol. 2017;149–150:1–20.

53. Frost JL, Schafer DP. Microglia: Architects of the Developing Nervous System. Trends Cell Biol. 2016;26(8):587–597.

54. Squarzoni P, et al. Microglia modulate wiring of the embryonic forebrain. Cell Rep. 2014;8(5):1271–1279.

55. Cunningham CL, Martínez-Cerdeño V, Noctor SC. Microglia regulate the number of neural precursor cells in the developing cerebral cortex. J Neurosci. 2013;33(10):4216–4233.

56. Hattori Y. The behavior and functions of embryonic microglia. Anat Sci Int. 2022;97(1):1–14.

57. Koyama R, Ikegaya Y. Microglia in the pathogenesis of autism spectrum disorders. Neurosci Res. 2015;100:1–5.

58. Zhan Y, et al. Deficient neuron-microglia signaling results in impaired functional brain connectivity and social behavior. Nat Neurosci. 2014;17(3):400–406.

59. Knuesel I, et al. Maternal immune activation and abnormal brain development across CNS disorders. Nat Rev Neurol. 2014;10(11):643–660.

60. Bian Z, et al. Deciphering human macrophage development at single-cell resolution. Nature. 2020;582(7813):571–576.

61. Askenase MH, et al. Longitudinal transcriptomics define the stages of myeloid activation in the living human brain after intracerebral hemorrhage. Sci Immunol. 2021;6(56). 10.1126/sciimmunol.abd6279

62. Sellgren CM, et al. Patient-specific models of microglia-mediated engulfment of synapses and neural progenitors. Mol Psychiatry. 2017;22(2):170–177.

63. Sellgren CM, et al. Increased synapse elimination by microglia in schizophrenia patient-derived models of synaptic pruning. Nat Neurosci. 2019;22(3):374–385.

64. Sheridan SD, et al. Umbilical cord blood-derived microglia-like cells to model COVID-19 exposure. Transl Psychiatry. 2021;11(1):179.

65. Satoh J-I, et al. TMEM119 marks a subset of microglia in the human brain. Neuropathology. 2016;36(1):39–49.

66. Stratoulias V, et al. Microglial subtypes: diversity within the microglial community. EMBO J. 2019;38(17):e101997.

67. Torres-Platas SG, et al. Morphometric characterization of microglial phenotypes in human cerebral cortex. J Neuroinflammation. 2014;11:12.

68. Tremblay M-È, et al. The role of microglia in the healthy brain. J Neurosci. 2011;31(45):16064–16069.

69. Leyh J, et al. Classification of Microglial Morphological Phenotypes Using Machine Learning. Front Cell Neurosci. 2021;15:701673.

70. Menassa DA, et al. The spatiotemporal dynamics of microglia across the human lifespan. Dev Cell. 2022;57(17):2127–2139.e6.

71. Sureshchandra S, et al. Single-cell RNA sequencing reveals immunological rewiring at the maternal-fetal interface following asymptomatic/mild SARS-CoV-2 infection. Cell Rep. 2022;39(11):110938.

72. Lu-Culligan A, et al. Maternal respiratory SARS-CoV-2 infection in pregnancy is associated with a robust inflammatory response at the maternal-fetal interface. Med (N Y). 2021;2(5):591–610.e10.

73. Argueta LB, et al. Inflammatory responses in the placenta upon SARS-CoV-2 infection late in pregnancy. iScience. 2022;25(5):104223.

74. Bordt EA, et al. Maternal SARS-CoV-2 infection elicits sexually dimorphic placental immune responses. Sci Transl Med. 2021;13(617):eabi7428.

75. Gao L, et al. Single-cell analysis reveals transcriptomic and epigenomic impacts on the maternal-fetal interface following SARS-CoV-2 infection. Nat Cell Biol. 2023;25(7):1047–1060.

76. Doratt BM, et al. Mild/Asymptomatic Maternal SARS-CoV-2 Infection Leads to Immune Paralysis in Fetal Circulation and Immune Dysregulation in Fetal-Placental Tissues. bioRxiv. [published online ahead of print: May 11, 2023]; 10.1101/2023.05.10.540233

77. Garcia-Flores V, et al. Maternal-fetal immune responses in pregnant women infected with SARS-CoV-2. Nat Commun. 2022;13(1):320.

78. Gomez-Lopez N, et al. Pregnancy-specific responses to COVID-19 revealed by high-throughput proteomics of human plasma. Commun Med. 2023;3(1):48.

79. Shook LL, et al. COVID-19 in pregnancy: implications for fetal brain development. Trends Mol Med. 2022;28(4):319–330.

80. Carbonnel M, et al. Plasticity of natural killer cells in pregnant patients infected with SARS-CoV-2 and their neonates during childbirth. Front Immunol. 2022;13:893450.

81. Foo S-S, et al. The systemic inflammatory landscape of COVID-19 in pregnancy: Extensive serum proteomic profiling of mother-infant dyads with in utero SARS-CoV-2. Cell Rep Med. 2021;2(11):100453.

82. Matute JD, et al. Single-cell immunophenotyping of the fetal immune response to maternal SARS-CoV-2 infection in late gestation. Pediatr Res. [published online ahead of print: November 8, 2021]; 10.1038/s41390-021-01793-z

83. Tang G, et al. Loss of mTOR-dependent macroautophagy causes autistic-like synaptic pruning deficits. Neuron. 2014;83(5):1131–1143.

84. Carter M, et al. Maternal Immune Activation and Interleukin 17A in the Pathogenesis of Autistic Spectrum Disorder and Why It Matters in the COVID-19 Era. Front Psychiatry. 2022;13:823096.

85. Malkova NV, et al. Maternal immune activation yields offspring displaying mouse versions of the three core symptoms of autism. Brain Behav Immun. 2012;26(4):607–616.

86. Gomez-Lopez N, et al. Distinct Cellular Immune Responses to SARS-CoV-2 in Pregnant Women. J Immunol. 2022;208(8):1857–1872.

87. Paolicelli RC, et al. Synaptic pruning by microglia is necessary for normal brain development. Science. 2011;333(6048):1456–1458.

88. Kim HJ, et al. Deficient autophagy in microglia impairs synaptic pruning and causes social behavioral defects. Mol Psychiatry. 2017;22(11):1576–1584.

89. Lenz KM, Nelson LH. Microglia and Beyond: Innate Immune Cells As Regulators of Brain Development and Behavioral Function. Front Immunol. 2018;9:698.

90. Baines KJ, West RC. Sex differences in innate and adaptive immunity impact fetal, placental, and maternal health. Biol Reprod. 2023;109(3):256–270.

91. Weinhard L, et al. Sexual dimorphism of microglia and synapses during mouse postnatal development: Sexual Dimorphism in Microglia and Synapses. Dev Neurobiol. 2018;78(6):618–626.

92. Bordt EA, et al. Isolation of Microglia from Mouse or Human Tissue. STAR Protoc. 2020;1(1). 10.1016/j.xpro.2020.100035

93. Wu T, et al. clusterProfiler 4.0: A universal enrichment tool for interpreting omics data. Innovation (Camb). 2021;2(3):100141.

94. Stirling DR, et al. CellProfiler 4: improvements in speed, utility and usability. BMC Bioinformatics. 2021;22(1):433.

