## Supplemental Figures and Table for "Maternal SARS-CoV-2 impacts fetal placental macrophage programs and placenta-derived microglial models of neurodevelopment"

**SUPPLEMENTAL INFORMATION**

**Figure S1.**

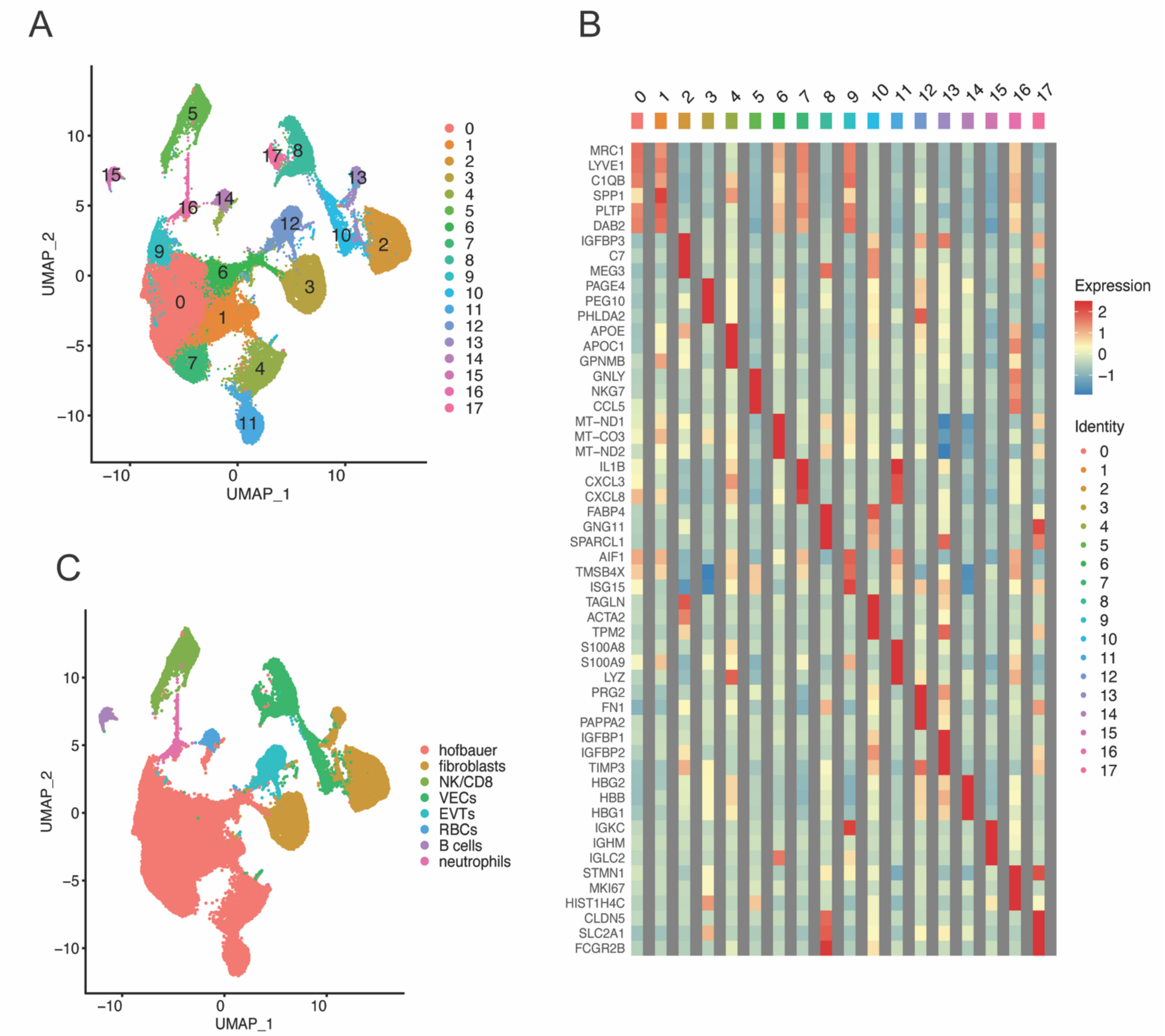


**Figure S1. Initial graph-based clustering and cluster assignments.** (**A**) Uniform Manifold Approximation and Projection (UMAP) visualization of 70,817 high-quality placental cells, enriched from placentas of pregnancies with (n=4) and without (n=8) SARS-CoV-2 infection, shows 18 clusters. (**B**) Heatmap displaying expression ($log_2$ fold change) of the top 3 marker genes per cluster. (**C**) UMAP visualization of cluster assignment by marker gene analysis demonstrating largest proportion of cells were macrophages/monocytes (labeled "hofbauer") and subset for further quality control and reclustering. NK: natural killer cells. VECs: vascular endothelial cells. EVTs: extravillous trophoblasts. RBCs: red blood cells.

**Figure S2.**

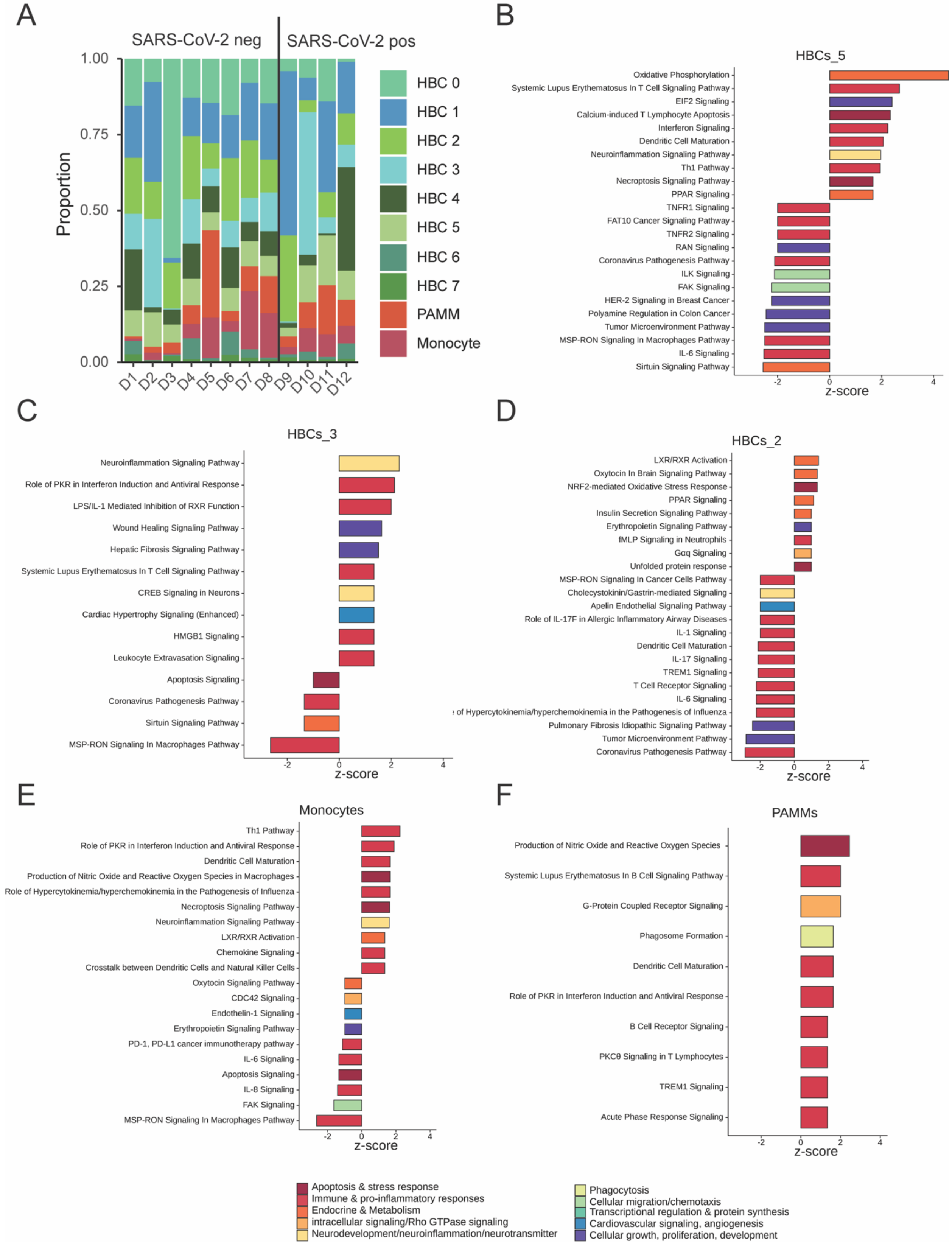
A
SARS-CoV-2 neg
SARS-CoV-2 pos
Proportion
1.00
0.75
0.50
0.25
0.00
D1 D2 D3 D4 D5 D6 D7 D8 D9 D10 D11 D12
HBC 0
HBC 1
HBC 2
HBC 3
HBC 4
HBC 5
HBC 6
HBC 7
PAMM
Monocyte
B
HBCs_5
Oxidative Phosphorylation
Systemic Lupus Erythematosus In T Cell Signaling Pathway
EIF2 Signaling
Calcium-induced T Lymphocyte Apoptosis
Interferon Signaling
Dendritic Cell Maturation
Neuroinflammation Signaling Pathway
Th1 Pathway
Necroptosis Signaling Pathway
PPAR Signaling
TNFR1 Signaling
FAT10 Cancer Signaling Pathway
TNFR2 Signaling
RAN Signaling
Coronavirus Pathogenesis Pathway
ILK Signaling
FAK Signaling
HER-2 Signaling in Breast Cancer
Polyamine Regulation in Colon Cancer
Tumor Microenvironment Pathway
MSP-RON Signaling In Macrophages Pathway
IL-6 Signaling
Sirtuin Signaling Pathway
-2 0 2 4
z-score
C
HBCs_3
Neuroinflammation Signaling Pathway
Role of PKR in Interferon Induction and Antiviral Response
LPS/IL-1 Mediated Inhibition of RXR Function
Wound Healing Signaling Pathway
Hepatic Fibrosis Signaling Pathway
Systemic Lupus Erythematosus In T Cell Signaling Pathway
CREB Signaling in Neurons
Cardiac Hypertrophy Signaling (Enhanced)
HMGB1 Signaling
Leukocyte Extravasation Signaling
Apoptosis Signaling
Coronavirus Pathogenesis Pathway
Sirtuin Signaling Pathway
MSP-RON Signaling In Macrophages Pathway
-2 0 2 4
z-score
D
HBCs_2
LXR/RXR Activation
Oxytocin In Brain Signaling Pathway
NRF2-mediated Oxidative Stress Response
PPAR Signaling
Insulin Secretion Signaling Pathway
Erythropoietin Signaling Pathway
fMLP Signaling in Neutrophils
Gαq Signaling
Unfolded protein response
MSP-RON Signaling In Cancer Cells Pathway
Cholecystokinin/Gastrin-mediated Signaling
Apelin Endothelial Signaling Pathway
Role of IL-17F in Allergic Inflammatory Airway Diseases
IL-1 Signaling
Dendritic Cell Maturation
IL-17 Signaling
TREM1 Signaling
T Cell Receptor Signaling
IL-6 Signaling
of Hypercytokinemia/hyperchemokinemia in the Pathogenesis of Influenza
Pulmonary Fibrosis Idiopathic Signaling Pathway
Tumor Microenvironment Pathway
Coronavirus Pathogenesis Pathway
-2 0 2 4
z-score
E
Monocytes
Th1 Pathway
Role of PKR in Interferon Induction and Antiviral Response
Dendritic Cell Maturation
Production of Nitric Oxide and Reactive Oxygen Species in Macrophages
Role of Hypercytokinemia/hyperchemokinemia in the Pathogenesis of Influenza
Necroptosis Signaling Pathway
Neuroinflammation Signaling Pathway
LXR/RXR Activation
Chemokine Signaling
Crosstalk between Dendritic Cells and Natural Killer Cells
Oxytocin Signaling Pathway
CDC42 Signaling
Endothelin-1 Signaling
Erythropoietin Signaling Pathway
PD-1, PD-L1 cancer immunotherapy pathway
IL-6 Signaling
Apoptosis Signaling
IL-8 Signaling
FAK Signaling
MSP-RON Signaling In Macrophages Pathway
-2 0 2 4
z-score
F
PAMMs
Production of Nitric Oxide and Reactive Oxygen Species
Systemic Lupus Erythematosus In B Cell Signaling Pathway
G-Protein Coupled Receptor Signaling
Phagosome Formation
Dendritic Cell Maturation
Role of PKR in Interferon Induction and Antiviral Response
B Cell Receptor Signaling
PKCθ Signaling in T Lymphocytes
TREM1 Signaling
Acute Phase Response Signaling
-2 0 2 4
z-score
Apoptosis & stress response
Immune & pro-inflammatory responses
Endocrine & Metabolism
intracellular signaling/Rho GTPase signaling
Neurodevelopment/neuroinflammation/neurotransmitter
Phagocytosis
Cellular migration/chemotaxis
Transcriptional regulation & protein synthesis
Cardiovascular signaling, angiogenesis
Cellular growth, proliferation, development

**Figure S2. Impact of maternal SARS-CoV-2 infection on Hofbauer cell subclusters, PAMMs and monocytes.** HBC: Hofbauer cell. PAMM: placenta-associated macrophage/monocyte. (**A**) Barplot demonstrating proportion of cells within each subcluster by participant. "SARS-CoV-2 neg" = no maternal SARS-CoV-2 infection during pregnancy. "SARS-CoV-2 pos" = positive maternal SARS-CoV-2 test during pregnancy. (**B-F**) Ingenuity Pathway Analysis (IPA) canonical pathways enrichment results of DEG by subcluster. IPA canonical pathways with absolute Z-score ≥1 and adjusted P-value $< 0.05$ are displayed.

**Figure S3.**

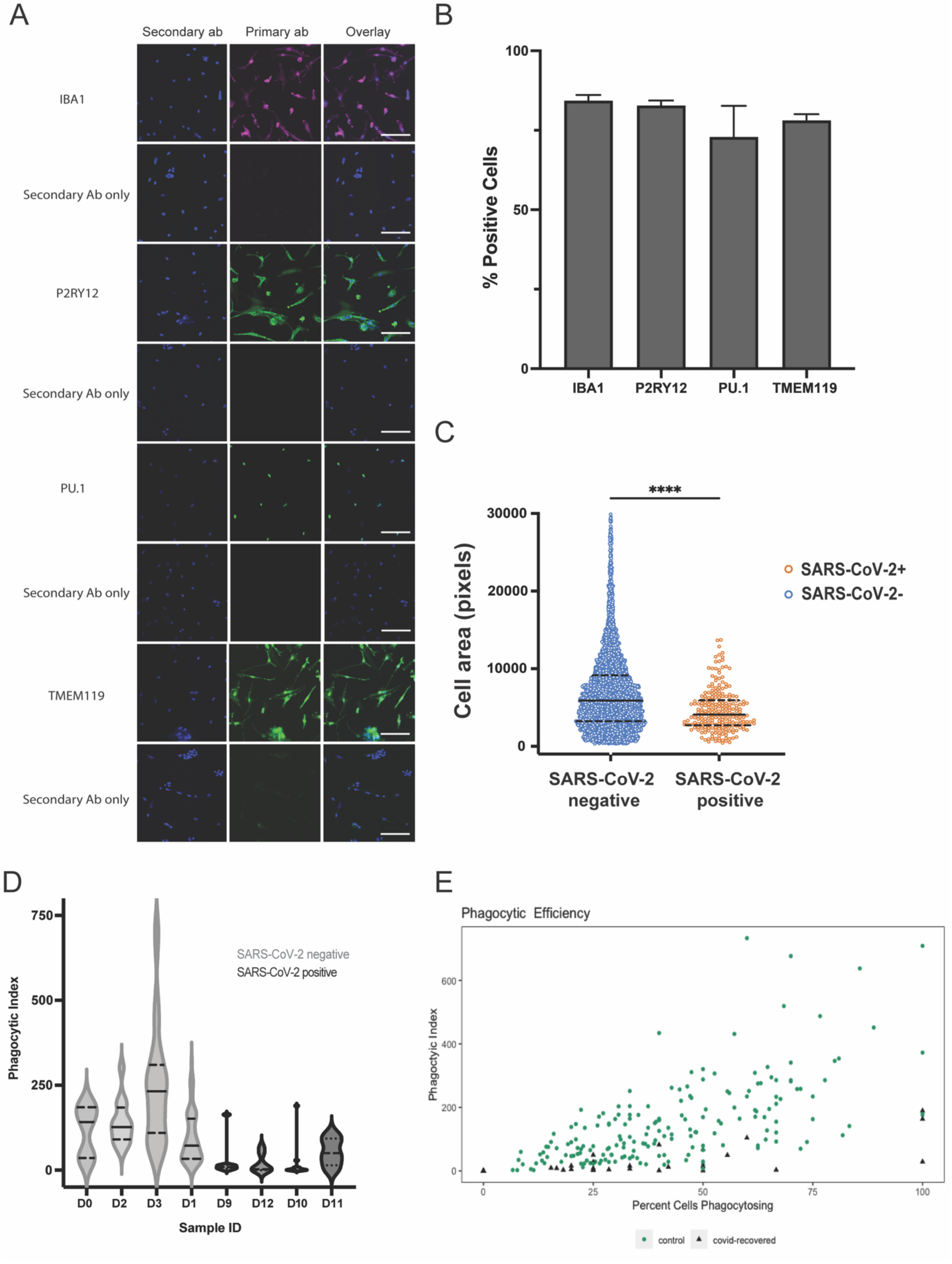
A
Secondary ab
Primary ab
Overlay
IBA1
Secondary Ab only
P2RY12
Secondary Ab only
PU.1
Secondary Ab only
TMEM119
Secondary Ab only
B
% Positive Cells
100
50
0
IBA1
P2RY12
PU.1
TMEM119
C
****
Cell area (pixels)
30000
20000
10000
0
SARS-CoV-2+
SARS-CoV-2-
SARS-CoV-2 negative
SARS-CoV-2 positive
D
Phagocytic Index
750
500
250
0
SARS-CoV-2 negative
SARS-CoV-2 positive
D0
D2
D3
D1
D9
D12
D10
D11
Sample ID
E
Phagocytic Efficiency
Phagocytic Index
600
400
200
0
0
25
50
75
100
Percent Cells Phagocytosing
control
covid-recovered

**Figure S3. HBC-iMG phenotypic and functional characterization.** HBC-iMGs: Hofbauer cells transdifferentiated toward microglia-like cells. Ab = antibody. (**A**) Confocal microscopy of immunostaining with primary antibody, secondary antibody (Hoechst nuclear stain) and overlay, showing lack of nonspecific staining. Scale bar = 100 μm. (**B**) Percentage of cells per sample with positive immunostaining. (**C**) Violin plots representing cell area (pixels) from cells in SARS-CoV-2 negative controls (blue, n=5257 cells) and positive cases (orange, n=232 cells). Solid lines represent median values and dashed lines interquartile range. Group differences assessed by Mann-Whitney U Test. ****$P<0.0001$. (**D**) Violin plot of phagocytic index per participant. Solid lines represent median values and dashed lines interquartile range. (**E**) Phagocytic Index by percent of cells engaged in any phagocytosis, by SARS-CoV-2 status. Green circles: SARS-CoV-2 negative controls (n=187 fields). Black triangles: SARS-CoV-2 positive cases (n=32 fields).

**Figure S4.**

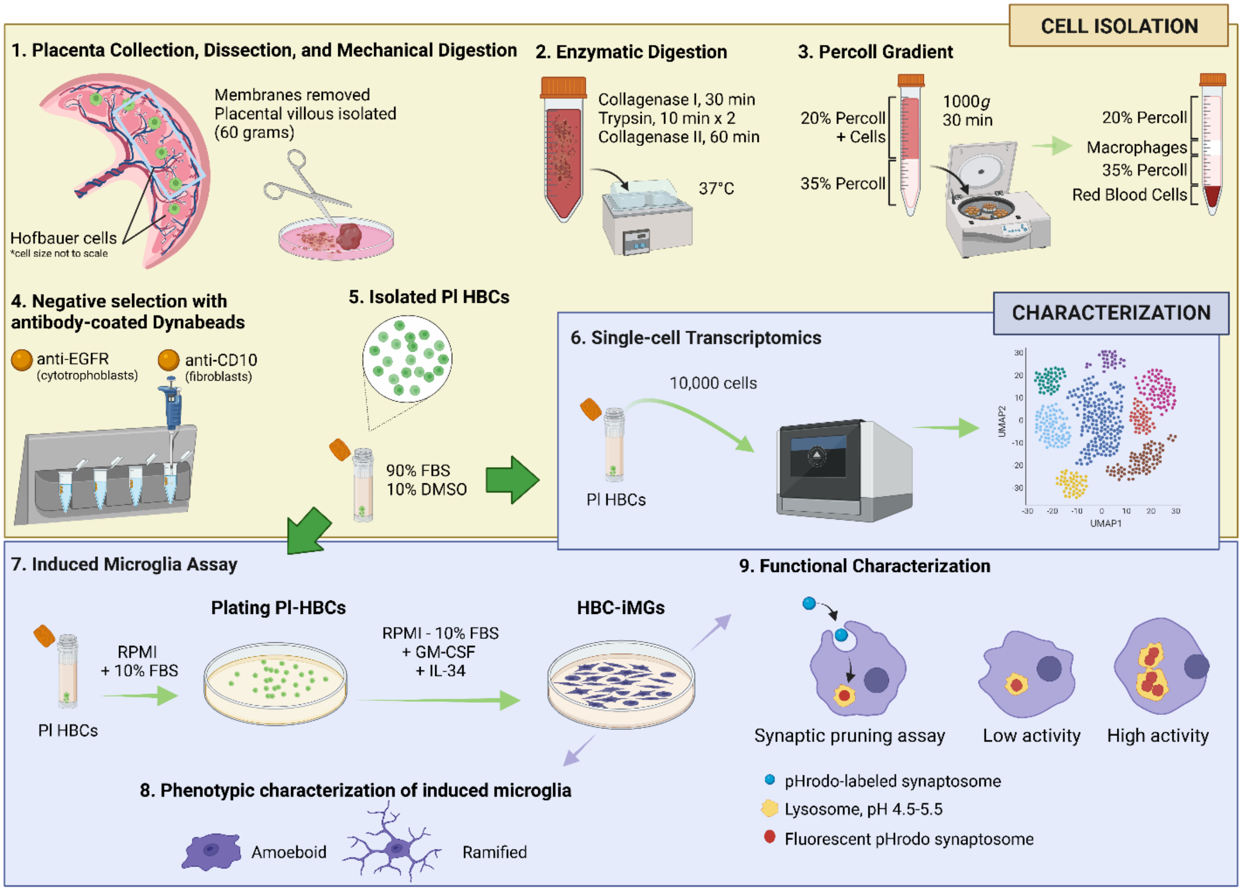


**Figure S4. Schematic of Hofbauer cell isolation procedure workflow and downstream analyses.** HBCs: Hofbauer cells. HBC-iMGs: Hofbauer cells transdifferentiated toward microglia-like cells. HBCs were isolated using collagenase and trypsin digestions, followed by centrifugation on Percoll gradients, and negative bead-based selection with antibody-coated anti-EGFR and anti-CD10 Dynabeads. HBCs were then characterized by single-cell RNA-Seq (10X Genomics) and cytokine induction to create HBC-iMGs. Image created in BioRender.

**Table S1. List of reagents used in Hofbauer cell isolation procedure.**

| Reagents | Source | Cat# |
|---|---|---|
| **Cytowash** | | |
| DMEM Medium | Thermo Fisher Scientific | 11965118 |
| Fetal Bovine Serum | Cytiva | SH 30071.03 |
| Pen/Strep (10,000 U/mL) | Thermo Fisher Scientific | 15140122 |
| Gentamicin | Thermo Fisher Scientific | 15750060 |
| **DPBS Wash** | | |
| 1X DPBS, Ca-/Mg-free | Thermo Fisher Scientific | 14190144 |
| Pen/Strep (10,000 U/mL) | Thermo Fisher Scientific | 15140122 |
| **Collagenase Digestion 1** | | |
| 1X DPBS, Ca-/Mg-free | Thermo Fisher Scientific | 14190144 |
| Collagenase Type IA | Millipore Sigma | 11088793001 |
| DNase I | Millipore Sigma | DN25-1G |
| Hyaluronidase | Millipore Sigma | H3506-1G |
| Bovine Serum Albumin | Millipore Sigma | A7906 |
| CaCl (3M stock, 1000x) | Boston Bio Products | MT-140-3M |
| **Trypsin Digestion** | | |
| 1X DPBS, Ca-/Mg-free | Thermo Fisher Scientific | 14190144 |
| Trypsin | Thermo Fisher Scientific | Gibco 27250018 |
| DNase I | Sigma Aldrich | DN25-1G |
| CaCl (3M stock, 1000x) | Boston Bio Products | MT-140-3M |
| **Collagenase Digestion 2** | | |
| 1X DPBS, Ca-/Mg-free | Thermo Fisher Scientific | 14190144 |
| Collagenase Type IA | Millipore Sigma | 11088793001 |
| DNase I | Millipore Sigma | DN25-1G |
| Hyaluronidase | Millipore Sigma | H3506-1G |
| Bovine Serum Albumin | Millipore Sigma | A7906 |
| CaCl (3M stock, 1000x) | Boston Bio Products | MT-140-3M |
| Dispase II | Thermo Fisher Scientific | 171105041 |
| **Gradient Solutions** | | |
| Percoll | GE Healthcare Biosciences | 17-0891-01 |
| Hanks' Balanced Salt Solution 10X | Thermo Fisher Scientific | 14185052 |
| Hanks' Balanced Salt Solution 1X | Thermo Fisher Scientific | 14175095 |
| OptiPrep Density Gradient Medium | Millipore Sigma | D1556-250mL |
| **Dynabeads** | | |
| Goat anti-mouse IgG antibody conjugated Dynabeads | Thermo Fisher Scientific | 11033 |
| Mouse anti-human EGFR | Thermo Fisher Scientific | MA5-16944 |
| Mouse anti-human CD10 antibody | Thermo Fisher Scientific | MA5-13070 |
